# Improving the Prediction of Clinical Success Using Machine Learning

**DOI:** 10.1101/2021.02.01.21250864

**Authors:** Bernard Munos, Jan Niederreiter, Massimo Riccaboni

**Author notes:** We thank Evaluate Ltd. for giving us access to the EvaluatePharma^®^ data used in this study and for their valuable feedback. The views expressed in this work are those of the authors. Prof. Riccaboni and Mr. Munos have been members of the Evaluate Ltd. “Forecasting Advisory Board” 2018-2019.

## Abstract

In pharmaceutical research, assessing drug candidates’ odds of success as they move through clinical research often relies on crude methods based on historical data. However, the rapid progress of machine learning offers a new tool to identify the more promising projects. To evaluate its usefulness, we trained and validated several machine learning algorithms on a large database of projects. Using various project descriptors as input data we were able to predict the clinical success and failure rates of projects with an average balanced accuracy of 83% to 89%, which compares favorably with the 56% to 70% balanced accuracy of the method based on historical data. We also identified the variables that contributed most to trial success and used the algorithm to predict the success (or failure) of assets currently in the industry pipeline. We conclude by discussing how pharmaceutical companies can use such model to improve the quantity and quality of their new drugs, and how the broad adoption of this technology could reduce the industry’s risk profile with important consequences for industry structure, R&D investment, and the cost of innovation

---

Machine learning (ML) tools are used with growing success across industries to improve decision-making. Businesses that have access to large volumes of high-quality data are increasingly turning to ML to perform tasks where it surpasses humans. From forecasting demand, resource needs, or financial performance; to predicting failure, detecting fraud, automating processes, reading X-rays; designing molecules; or understanding customer behavior, there is hardly a facet of business that cannot benefit from ML^1^. In the pharmaceutical industry, which spends more than $180 billion annually in research and development (R&D)^2^ but faces failure rates that often exceed 90%^3^, a model that could predict the outcomes of clinical research phases would be particularly valuable. Several pioneering contributions have already used ML to mine clinical trials data in order to predict the likelihood of trial success and regulatory approval for drug candidates^4,5,6^. Our paper extends this work by recognizing that clinical and regulatory success depend upon the complex interaction of a broad set of predictors that includes both trial-related variables as well as other success factors such as molecule attributes, regulatory status, patent protection, company features, and market data. To model these complex dynamics, we applied eight ML approaches to our data, which produced a best-performing algorithm (BART) that has never been used in this context. We also identified new, highly relevant predictors of success.

Section 2 below describes our ML methodology and dataset. Section 3 compares the performance of our “best-in-class” ML algorithm to the methods commonly used in industry. Section 4 illustrates one use of our ML approach by predicting the outcomes of the current industry pipeline. We conclude by summarizing our findings and discussing their potential implications for the pharmaceutical industry and biomedical research.

## 2. Data and Methods

**Box 1 | Machine Learning**

Learning computer algorithms, that evaluate and automatically improve their performance, go back many decades. In 1952 Arthur Samuel designed one of the first computer learning programs that improved its ability to play checkers by learning from previous moves. He coined the term “machine learning” (ML), which has come to designate a computer algorithm that ‘learns’ to better its performance on a specific task. Since the 1950s machine learning has made huge advances that have heightened its performance and broadened its appeal. Image recognition software, email filters, and personalized advertisement are just some of the applications which rely on ML technology. And thanks to the growing availability of large datasets, machine-learning is making its way into healthcare, including drug discovery^7^, medical imaging^8^, and health monitoring^8^.

A simple three-step machine-learning routine is depicted in Exhibit 1. An input dataset for which the outcome of interest is known, is randomly split into two subsets (step 1): a training set and a validation set. During the learning process the ML algorithm repeatedly evaluates pairs of input/output data from the training set (step 2). For each pair, it estimates an output value, and compares it to the true (known) value. The distance between the two is then used by the algorithm to fine-tune itself and improve its performance. As the training progresses, that distance shrinks, until it is consistently smaller than a pre-set value. At that point, the algorithm is deemed to be trained. (Note: if the input data do not have enough explanatory power, the training may fail, which is a signal that another, more accurate model is needed.)

**Exhibit 1:**
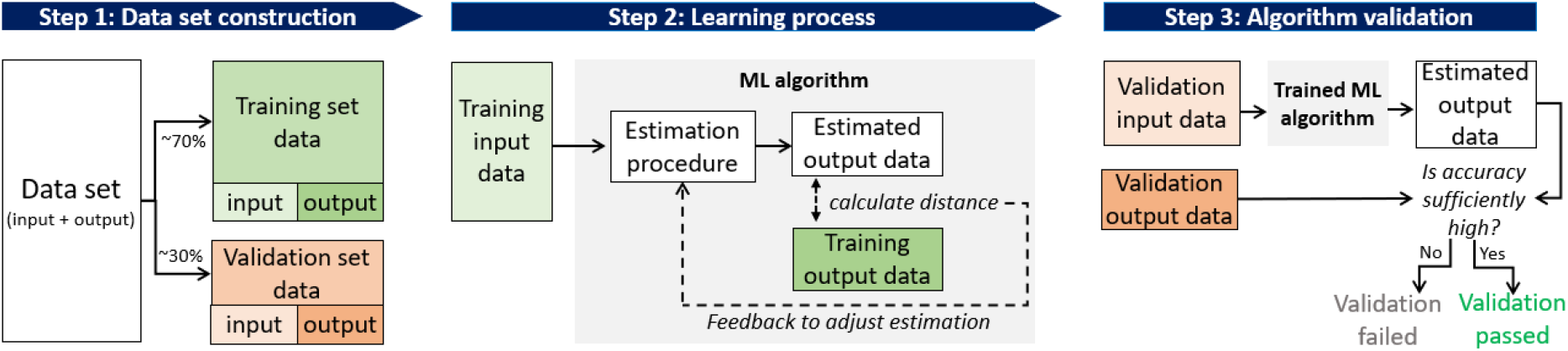
Example of a supervised machine learning routine.

Once it is trained, the algorithm is validated by applying it to the validation set – which it has never seen (step 3). To be successfully validated, it must estimate the output values of the validation set with an accuracy that is sufficient for its purpose. (Note: It is possible for the algorithm to fail the validation step. This can happen, for instance, when the input dataset is too small, causing the algorithm to “over-learn” the results, instead of predicting them.)

When the output data is binary (e.g. pass/fail) the performance of the algorithm can be summarized by a “confusion matrix” which relates classified successes and failures to true successes and failures. From the entries of the confusion matrix various performance measures can be derived that summarize the goodness-of-fit of the classification (see template in Exhibit 2). In this paper, we focus particularly on the area under the receiver operating curve (AUC)^9^ and balanced accuracy (BACC) which are widely used to assess the performance of classifiers.

**Exhibit 2:**
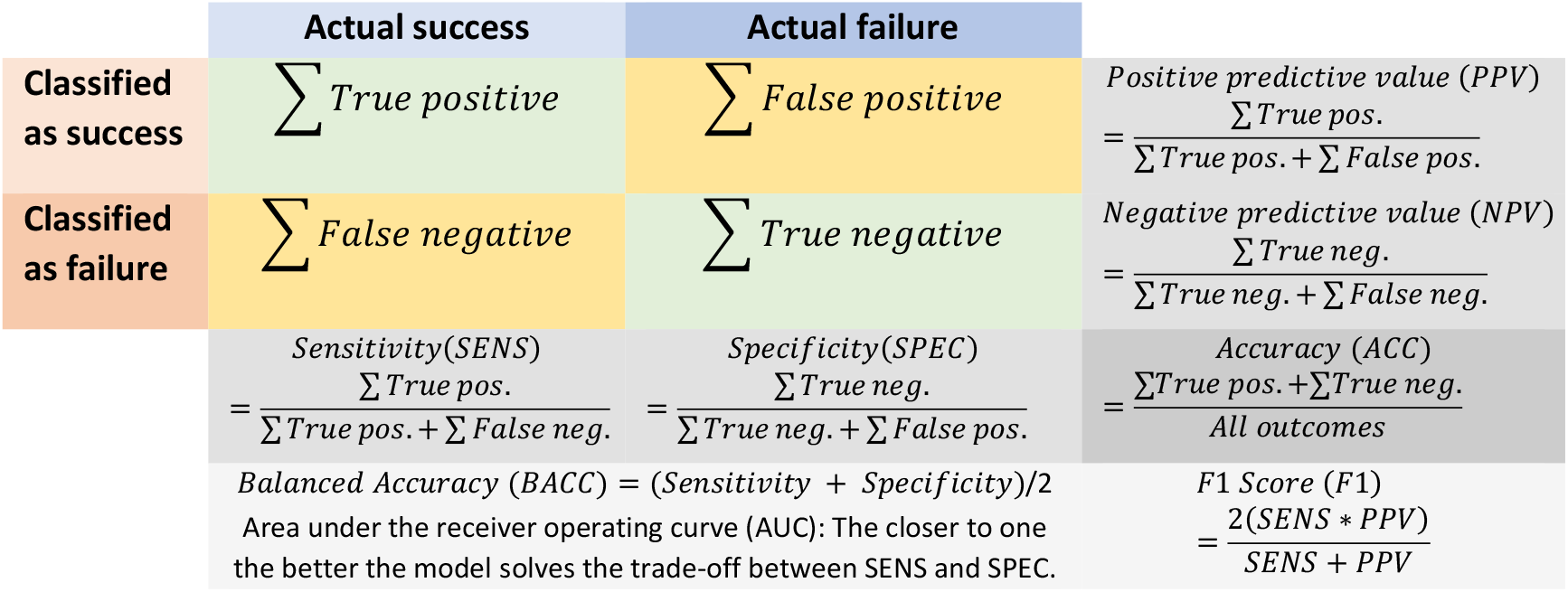
Confusion matrix template including performance measures.

After successful validation, the algorithm can be applied to similar, new input data and used to predict their (unknown) output. The great advantage of ML over traditional statistical methods such as regression or discriminant analyses, is that ML excels at modeling non-linear relationships (e.g., synergies and multiple feedback loops). Given such data, its performance is consistently better, as our example will illustrate.

We trained various algorithms on a database of drug development projects to predict the success or failure of the clinical research phases in which they were engaged. Each project is a combination of input and output data. The input data recapitulates the attributes of each project -- e.g., the features of the molecule; intended market; company; etc. – while the output data indicates the status of its most advanced clinical research phase – e.g., success, failure, or on-going. For instance, a project might be lorlatinib to treat ALK positive, non-small cell lung cancer. The input data would describe a small molecule developed by Pfizer that was granted expedited FDA review^a^ (see Exhibit 10 in the supplementary material A.1 for a detailed description of all features). The output data would indicate: “NDA/BLA & Approved or Marketed”^b^.

**Exhibit 10:**
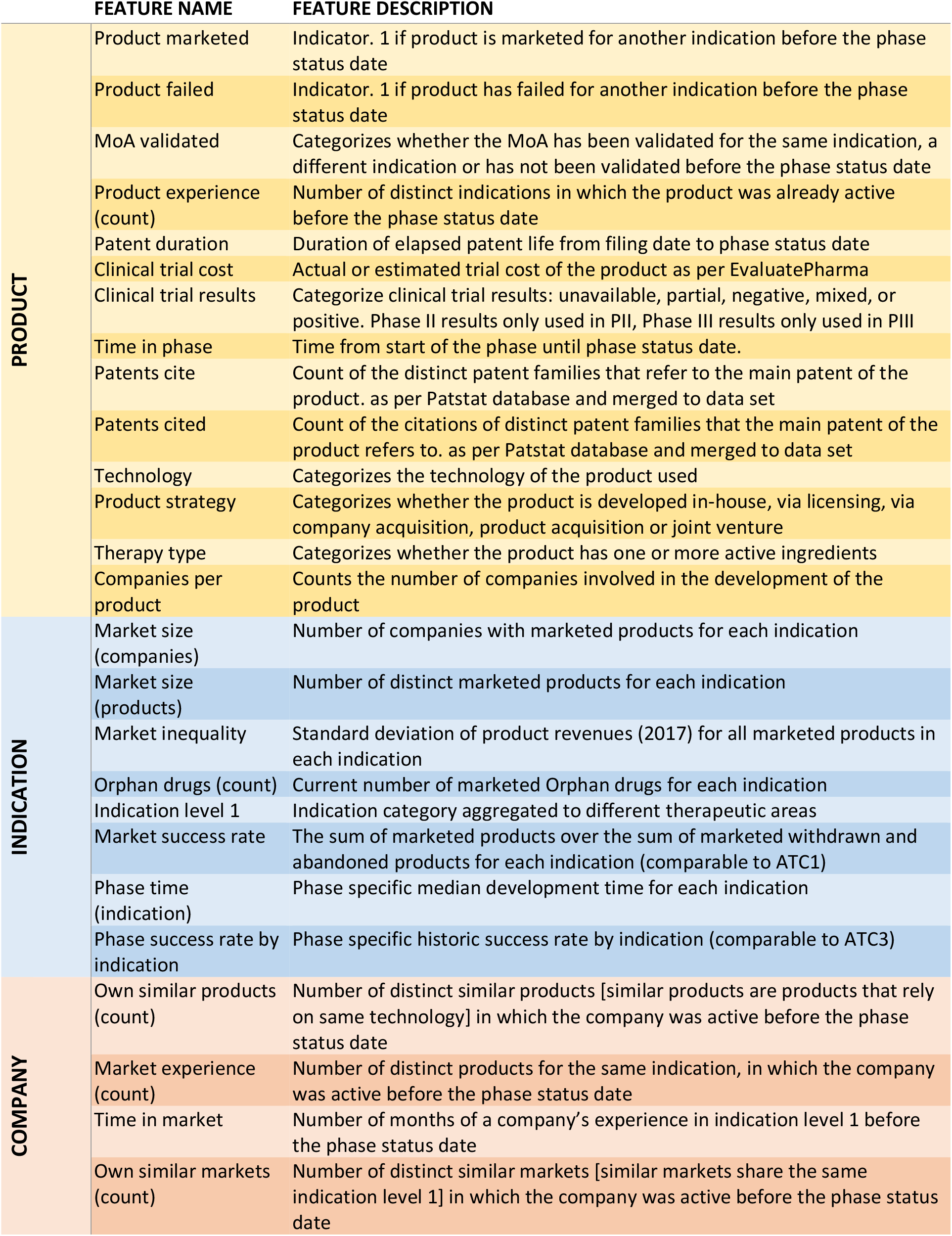
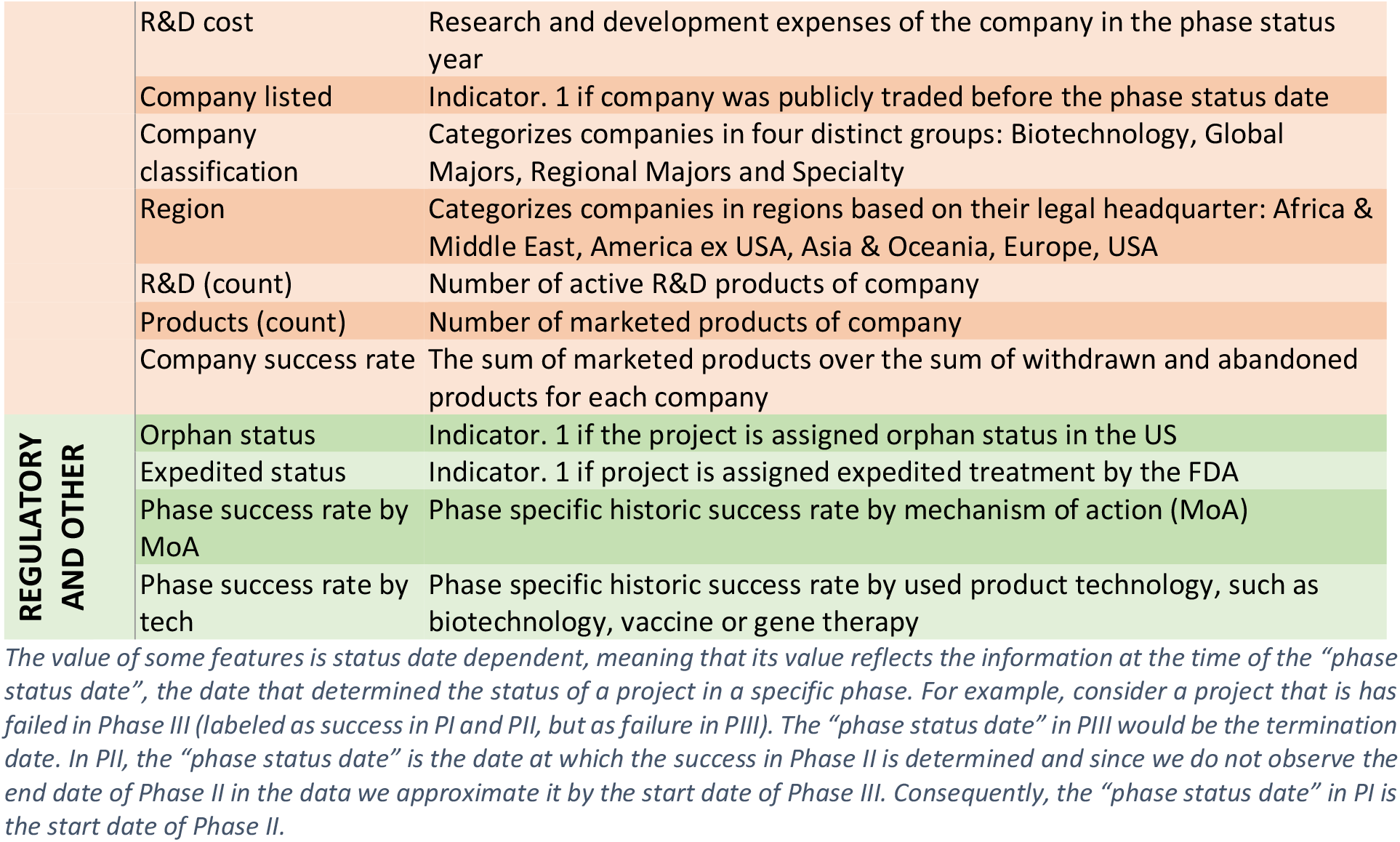
description of candidate features used in ML algorithms

Our data comes from a novel database created by Evaluate Ltd. to which we were granted access^c^. It includes 8,785 projects that were undertaken in the United States during the last decade. They encompass more than 4,500 NMEs and 1,300 companies and cover a wide range of indications. The anonymized dataset can be downloaded from supplementary material B, which includes a link to the executable R code.

The database was partitioned into three subsets (PI, PII, and PIII), for projects having reached phase I, II, and III respectively. For each subset, we randomly split the projects for which the outcome is known (i.e. failure or success) into a training and a validation set. Exhibit 3 shows the grid used to ascertain the output value of each project. It also shows the clinical success rates achieved for each phase by the molecules in our sample. Before training the ML algorithms, we evaluated different pre-processing techniques such as feature selection methods and various ways to deal with missing information.

**Exhibit 3:** Project status classification and number of projects for each clinical research phase

|  | Data sets according to clinical research phases |  |  |
| --- | --- | --- | --- |
| Project status | Phase I | Phase II | Phase III |
| **NDA/BLA & Approved or Marketed** | Success 498 | Success 499 | Success 579 |
| **Phase III, on-going** | Success 336 | Success 347 | On-going 559 |
| **Phase III, abandoned/suspended** | Success 248 | Success 147 | Failure 290 |
| **Phase II, on-going** | Success 844 | On-going 2372 |  |
| **Phase II, abandoned/suspended** | Success 735 | Failure 1794 |  |
| **Phase I, on-going** | On-going 1858 |  |  |
| **Phase I, abandoned/suspended** | Failure 1231 |  |  |
| **Total number of projects by phase\*** | **5750** | **5159** | **1428** |
| **Avg. success rate (success/ sum)** | **68.4%** | **35.7%** | **66.6%** |

*Projects that refer to combined phases are assigned to the earlier phase (e.g. phase2/3 is assigned to phase 2). The same project can be assigned to more than one dataset. For instance, a phase III project can be categorized as on-going in PIII, and successful in PI and PII. The total number of distinct projects as found in the database is 8785. Projects that have succeeded in phase III have been lumped into a “NDA/BLA & Approved or Marketed” category, as there were too few NDA/BLA projects to train algorithms, if that category was split out. (In our data only 2.6% fail between NDA/BLA and approval and 0.1% are not marketed even though approved), which results in too few observations to successfully train ML algorithms).*

After preprocessing, the training sets were used to train eight different ML algorithms: Bayesian additive regression tree (BART), random forest (RF), boosted decision trees (C5.0), support vector machine (SVM), probabilistic regression (PROBIT), artificial neural net (ANN), a simple decision tree (DT) and an ensemble learner, which were then applied to their respective validation sets. The results were compared using several performance metrics such as the area under the curve (AUC) and balanced accuracy (BACC, see Exhibit 2). The best performing algorithm across data sets – PI, PII and PIII – was referred to as “best-in-class”. Details about the pre-processing step and the training of the eight algorithms can be found in the supplementary material A.2.

In the next section, the “best-in-class” algorithm is compared to two common prediction methods – one based on historical data, and the other on discriminant analysis, which is frequently used to classify binary outcomes (success/failure).

## 3. Machine learning vs. common estimation methods

The Bayesian Additive Regression Tree (BART) method^10^ produced the best-performing algorithm for each dataset across to various performance measures (Exhibit 16 in the supplementary material A.2 contains the performance measures of each method on each data set). To add perspective, this section compares the BART results to the cruder method based on historic success rates and to discriminant analysis.

**Exhibit 16:** Average validation results across ML algorithms and data sets

| ML Method | Data set | AUC Mean | AUCL Mean | AUCH Mean | SENS Mean | SPEC Mean | PPV Mean | NPV Mean | F1 Mean | BACC Mean |
| --- | --- | --- | --- | --- | --- | --- | --- | --- | --- | --- |
| BART | PI | **0.93** | **0.92** | **0.95** | 0.91 | **0.76** | **0.89** | 0.80 | **0.90** | **0.83** |
|  | PII | **0.96** | **0.95** | **0.97** | **0.82** | **0.95** | **0.90** | **0.91** | **0.86** | **0.89** |
|  | PIII | **0.94** | **0.92** | **0.97** | 0.88 | **0.84** | **0.92** | 0.78 | **0.90** | **0.86** |
| RF | PI | 0.88 | 0.86 | 0.90 | **0.95** | 0.55 | 0.82 | **0.84** | 0.88 | 0.75 |
|  | PII | 0.93 | 0.91 | 0.95 | 0.81 | 0.90 | 0.82 | 0.89 | 0.81 | 0.85 |
|  | PIII | 0.91 | 0.87 | 0.94 | **0.94** | 0.61 | 0.83 | **0.83** | 0.88 | 0.77 |
| C 5.0 | PI | 0.86 | 0.84 | 0.89 | 0.92 | 0.58 | 0.83 | 0.76 | 0.87 | 0.75 |
|  | PII | 0.91 | 0.89 | 0.93 | 0.73 | 0.92 | 0.83 | 0.86 | 0.77 | 0.82 |
|  | PIII | 0.88 | 0.84 | 0.92 | 0.88 | 0.67 | 0.85 | 0.74 | 0.86 | 0.78 |
| SVM | PI | 0.86 | 0.83 | 0.88 | 0.92 | 0.53 | 0.81 | 0.76 | 0.86 | 0.72 |
|  | PII | 0.90 | 0.87 | 0.92 | 0.67 | 0.93 | 0.84 | 0.84 | 0.74 | 0.80 |
|  | PIII | 0.87 | 0.82 | 0.91 | 0.91 | 0.61 | 0.83 | 0.77 | 0.86 | 0.76 |
| PROBIT | PI | 0.84 | 0.82 | 0.87 | 0.90 | 0.56 | 0.82 | 0.72 | 0.86 | 0.73 |
|  | PII | 0.89 | 0.86 | 0.91 | 0.67 | 0.91 | 0.80 | 0.84 | 0.73 | 0.79 |
|  | PIII | 0.83 | 0.78 | 0.88 | 0.88 | 0.55 | 0.80 | 0.71 | 0.83 | 0.71 |
| ANN | PI | 0.75 | 0.73 | 0.78 | 0.88 | 0.54 | 0.81 | 0.62 | 0.84 | 0.71 |
|  | PII | 0.77 | 0.74 | 0.80 | 0.53 | 0.90 | 0.74 | 0.78 | 0.60 | 0.72 |
|  | PIII | 0.82 | 0.77 | 0.87 | 0.88 | 0.59 | 0.82 | 0.70 | 0.84 | 0.73 |
| DT | PI | 0.69 | 0.66 | 0.72 | 0.92 | 0.44 | 0.78 | 0.71 | 0.84 | 0.68 |
|  | PII | 0.85 | 0.82 | 0.88 | 0.60 | 0.91 | 0.78 | 0.80 | 0.67 | 0.75 |
|  | PIII | 0.79 | 0.73 | 0.85 | 0.88 | 0.58 | 0.81 | 0.71 | 0.84 | 0.73 |
| Ensemble learner | PI | 0.93 | 0.91 | 0.95 | 0.92 | 0.72 | 0.87 | 0.82 | 0.89 | 0.82 |
|  | PII | 0.91 | 0.88 | 0.94 | 0.84 | 0.85 | 0.75 | 0.91 | 0.79 | 0.85 |
|  | PIII | 0.97 | 0.94 | 0.99 | 0.97 | 0.70 | 0.82 | 0.95 | 0.89 | 0.84 |

Abbreviations: AUC - Area under the receiver operating characteristic curve; AUCL(H) - Lower (Upper) 95% AUC confidence interval calculated for each repetition based on DeLong method<sup>11</sup>; SENS: Sensitivity; SPEC: Specificity; PPV: Positive predictive value, NPV: Negative predictive value; F1: F1 Score; BACC: Balanced accuracy. Highlighted in bold are the values that correspond to the best performing algorithm for each phase.

The historical (HIST) method classifies projects as successful if the historic success rate for compounds targeting the same indication in the same phase is greater than 50%. The discriminant analysis (DISCR) is an adaptation of regression analysis to situations in which the dependent variable is qualitative (e.g., success vs. failures).

The results of this comparison are reported in Exhibit 4 which displays the average performance measures on the validation sets obtained from resampling randomly training and validation sets 100 times. They show that the ‘best-in-class’, BART algorithm classifies the outcomes of clinical research phases (e.g., success or failures) with a balanced accuracy of at least 83% (PI = 83%; PII = 89%; PIII = 86%). The AUC reaches 93%, 96%, and 94% for PI, PII and PIII respectively. The HIST method is markedly less accurate (BACC: PI = 56%; PII = 60%; PIII = 70%, AUC: PI=64%, PII=69%, PIII=79%). The DISCR performs better than HIST but is still significantly less accurate than the BART ML approach (BACC: PI = 73%; PII = 78%; PIII = 73%, AUC: PI=85%, PII=88%, PIII=84%).

**Exhibit 4:** Comparative performance of ‘best-in-class’ BART ML, historical method, and discriminant analysis

| ML Method | Data set | AUC Mean | AUCL Mean | AUCH Mean | SENS Mean | SPEC Mean | PPV Mean | NPV Mean | F1 Mean | BACC Mean |
| --- | --- | --- | --- | --- | --- | --- | --- | --- | --- | --- |
| BART | PI | 0.93 | 0.92 | 0.95 | 0.91 | 0.76 | 0.89 | 0.80 | 0.90 | 0.83 |
|  | PII | 0.96 | 0.95 | 0.97 | 0.82 | 0.95 | 0.90 | 0.91 | 0.86 | 0.89 |
|  | PIII | 0.94 | 0.92 | 0.97 | 0.88 | 0.84 | 0.92 | 0.78 | 0.90 | 0.86 |
| HIST | PI | 0.64 | 0.60 | 0.67 | 0.83 | 0.29 | 0.71 | 0.45 | 0.77 | 0.56 |
|  | PII | 0.69 | 0.65 | 0.74 | 0.26 | 0.94 | 0.70 | 0.71 | 0.38 | 0.60 |
|  | PIII | 0.79 | 0.73 | 0.86 | 0.71 | 0.70 | 0.82 | 0.55 | 0.76 | 0.70 |
| DISCR | PI | 0.85 | 0.79 | 0.85 | 0.89 | 0.56 | 0.82 | 0.71 | 0.85 | 0.73 |
|  | PII | 0.88 | 0.84 | 0.90 | 0.65 | 0.91 | 0.79 | 0.83 | 0.71 | 0.78 |
|  | PIII | 0.84 | 0.75 | 0.88 | 0.87 | 0.60 | 0.82 | 0.69 | 0.84 | 0.73 |

*Performance values are averaged over 100 repetitions for which the training/validation routine is performed. Abbreviations: AUC - Area under the receiver operating characteristic curve; AUCL(H) - Lower (Upper) 95% AUC confidence interval calculated for each repetition based on DeLong method<sup>11</sup>; SENS: Sensitivity; SPEC: Specificity; PPV: Positive predictive value, NPV: Negative predictive value; F1: F1 Score; BACC: Balanced accuracy.*

The differences between the ML and HIST methods are visualized in Exhibit 5. Across clinical phases, ML-predicted successes and failures appear more representative of their actual distributions than the predictions based on historic success rates. The same is true of the mean classification value (see Exhibit 23 in the supplementary material A.4 for a comparison between ML and DISCR).

**Exhibit 5:**
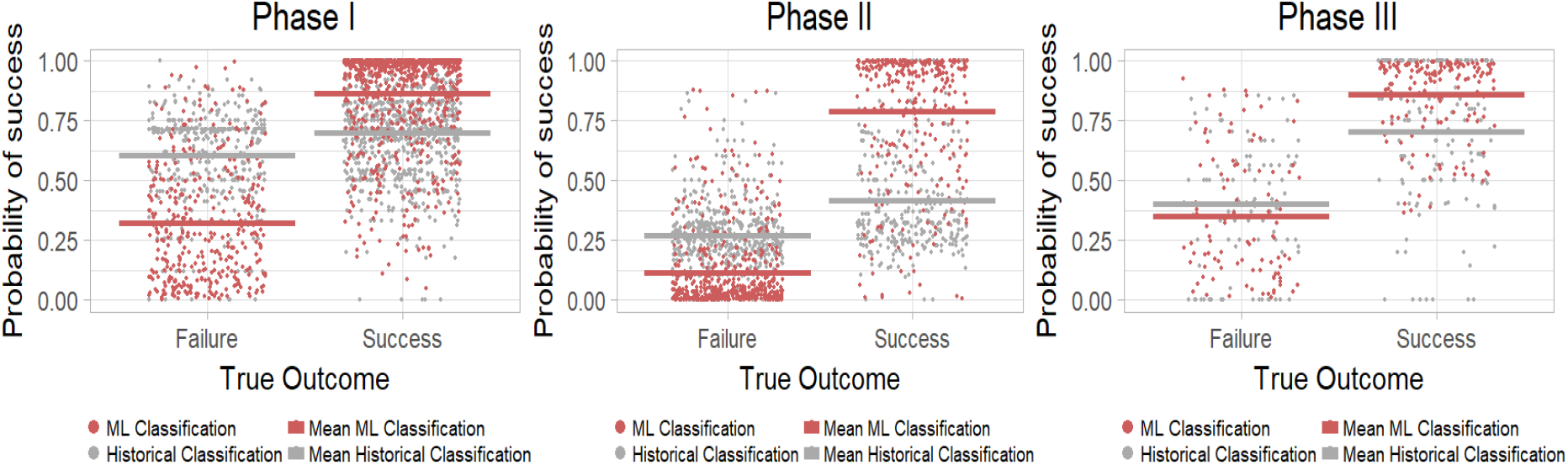
Best-in-class ML and historical classification values separated by phase and true outcome. The Exhibit shows for each phase the estimated success probabilities for failed and successful projects in the validation set using the BART method (red dots) and the historical method (grey dots). For each method, the estimated average success probability is depicted by horizontal lines. On average, the estimated success probability of successful (failed) projects is higher (lower) with BART than HIST.

**Exhibit 7:**
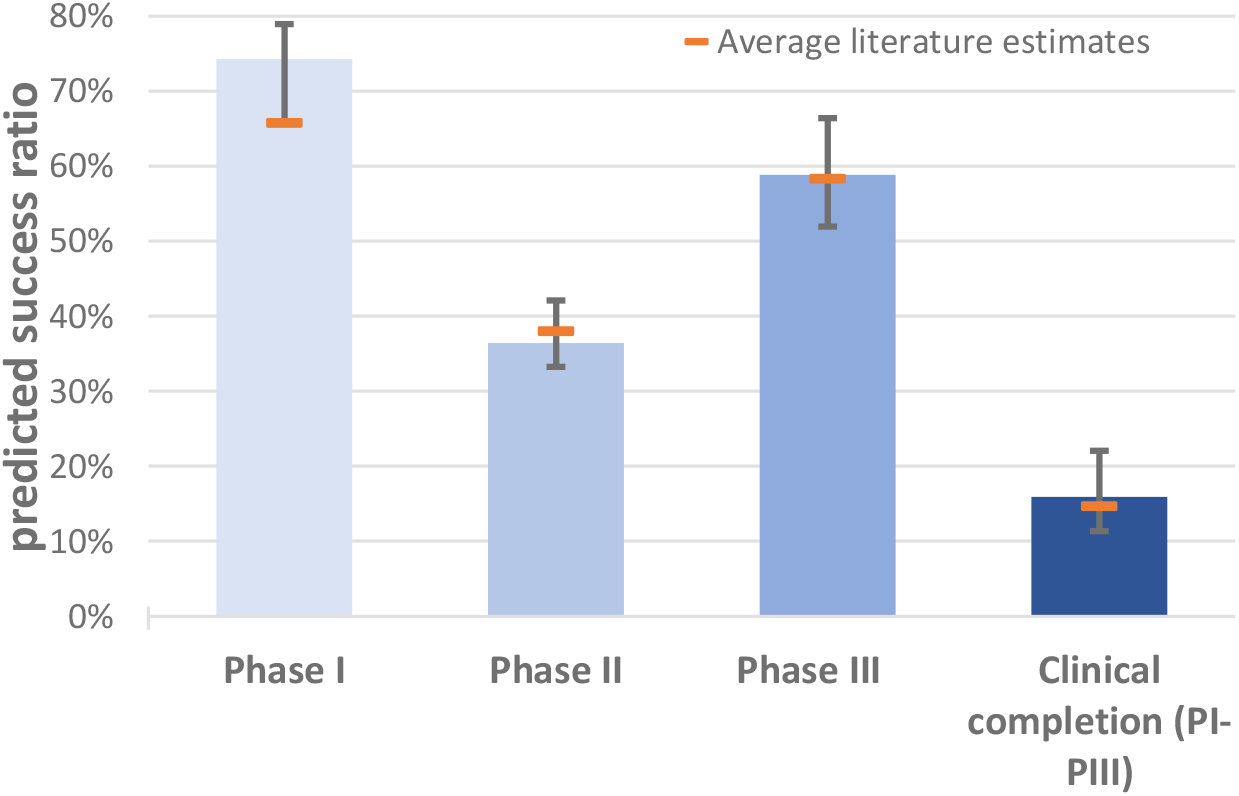
Predicted success rates of current project pipeline. The Exhibit shows the ML success ratio predicted for current pipeline projects split by phase and compares them to an average over literature estimates.

**Exhibit 8:**
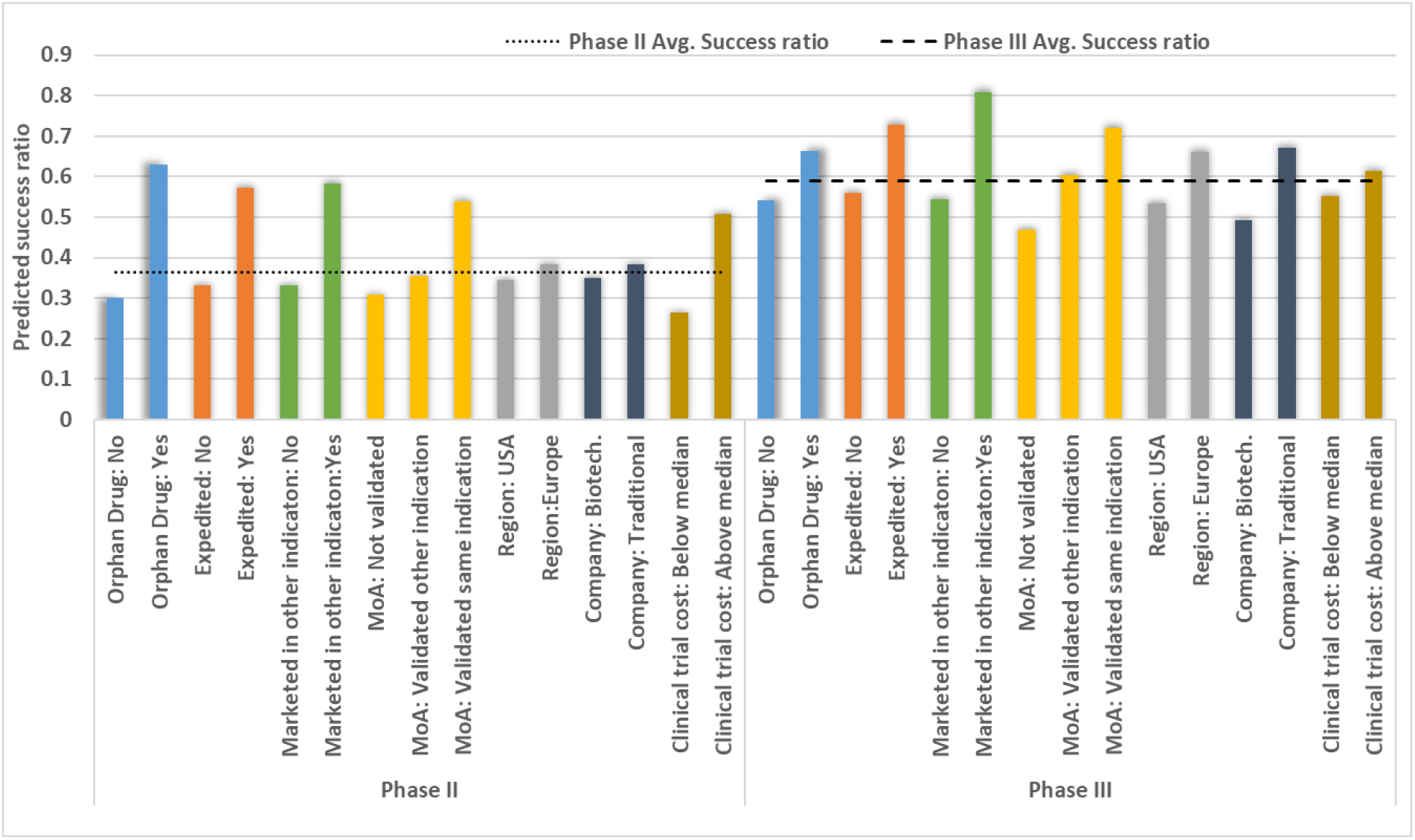
Predicted success rate of current PII/PIII pipeline by success factors. The Exhibit shows the success ratio predicted of current Phase II and III pipeline projects split various categories. The average predicted success ratios across projects are depicted by horizontal lines. The numbers in the text refer to the difference between predicted success ratio of a category and average predicted success ratio.

**Exhibit 11:**
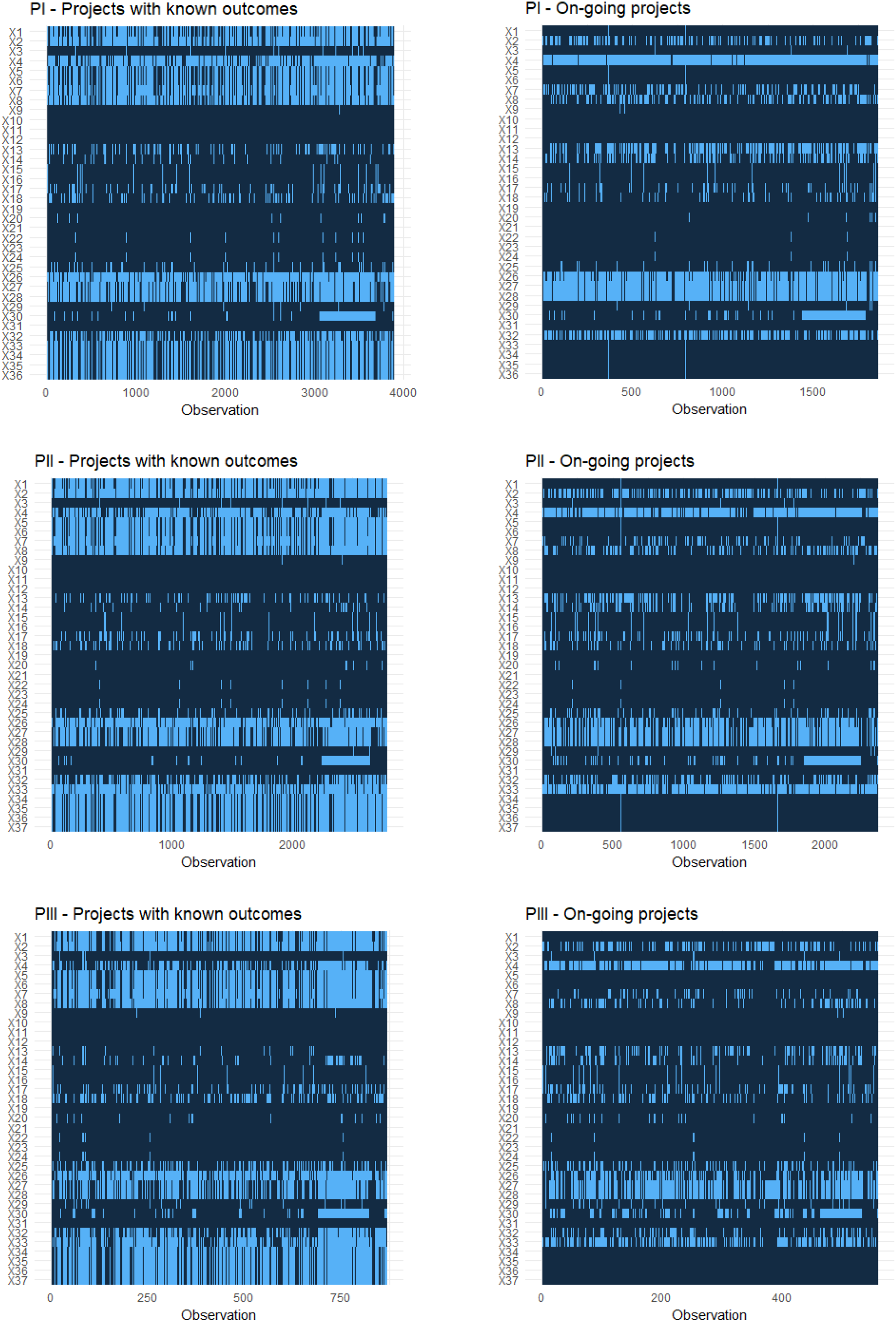
Missingness analysis (missing features across data sets) The graphs visualize the missing features (light blue) for each observation in the data sets PI, PII and PIII are split according to whether the outcome of a project is known or is on-going.

**Exhibit 12:**
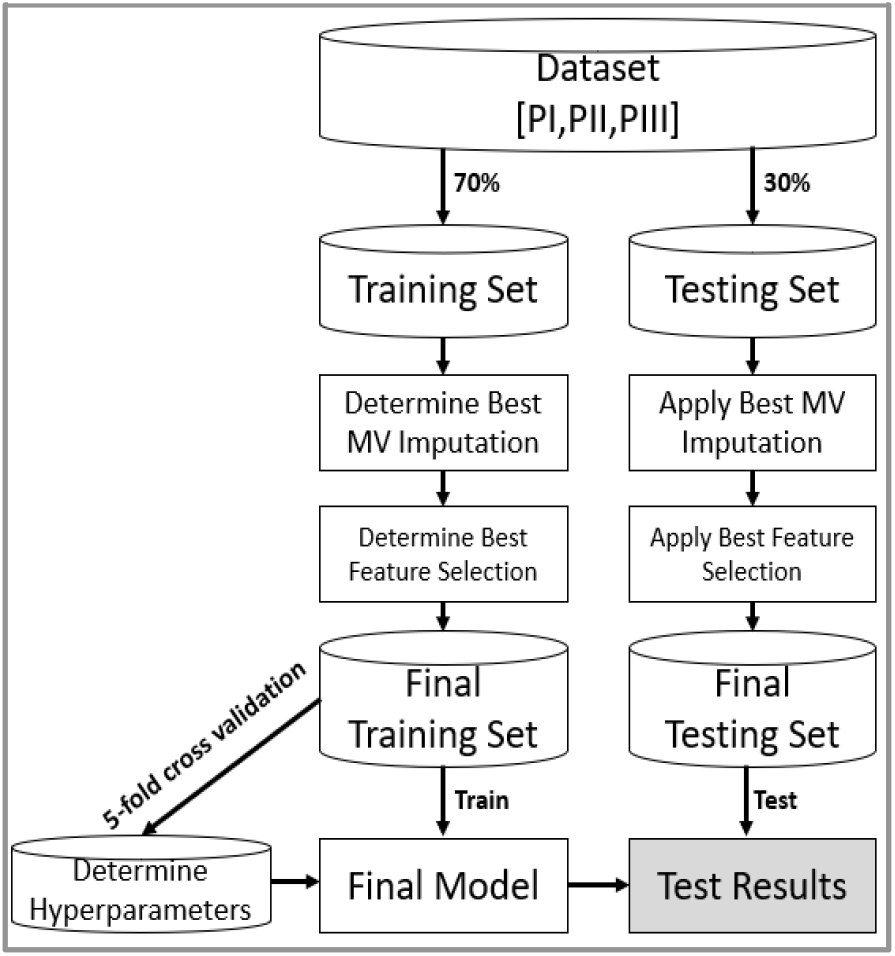
Applied training and testing routine.

**Exhibit 23:**
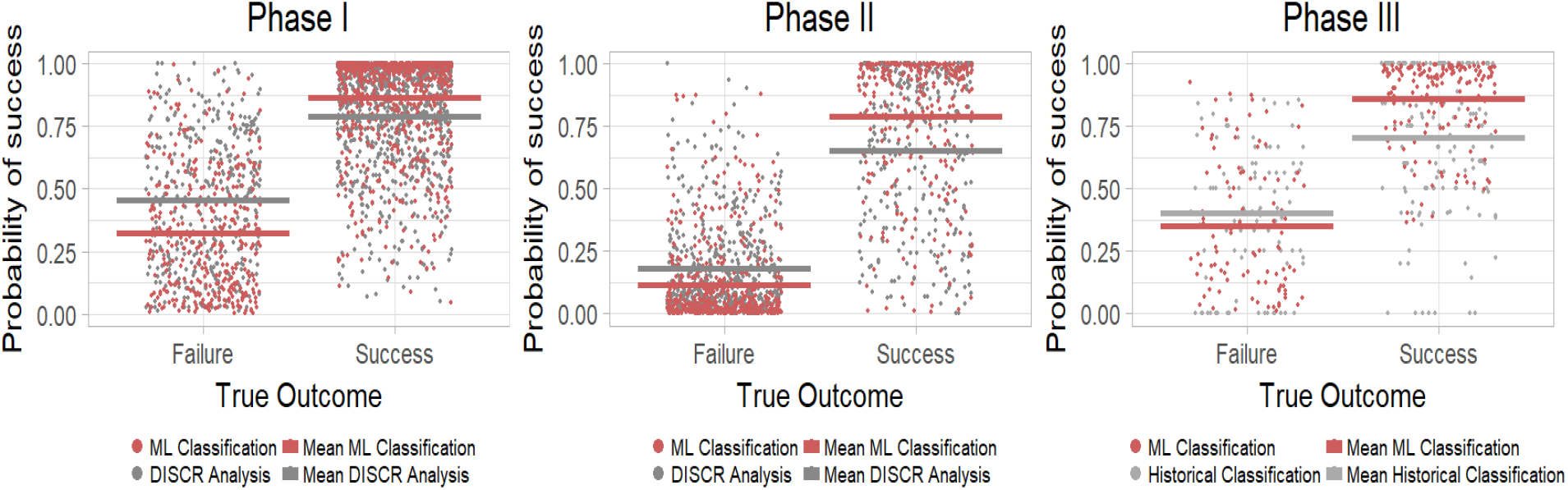
ML and discriminant analysis classification values separated by phase and true outcome. The Exhibit shows for each phase the estimated success probabilities for failed and successful projects in the validation set using the BART method (red dots) and the discriminant method (grey dots). For each category, the estimated average success probability is depicted by horizontal lines. On average, the estimated success probability of successful (failed) projects is estimated higher (lower) using BART in comparison to DISCR.

The better performance of the BART algorithm relative to discriminant analysis derives from its ability to handle missing information in the input dataset^12^; to include features based on their contribution to performance^13^; and to exploit hidden relations between project features. Analyzing the features that are most prominently selected during the training phase can point us to the kind of information is useful to boost predictive performance. We found that the features most frequently selected by the algorithm across phases relate to company, product, market and regulatory status. Exhibit 6 shows that information on company, indication, market, and mode of action (MoA) success rates as well as clinical trial costs, patent duration and expedited FDA review is frequently selected by the algorithm and interactions across features are common^d^.

**Exhibit 6:** Most relevant features in ‘best-in-class’ algorithm

| Feature Importance by singular features |  |  |  |  |  |  |  |  |  |
| --- | --- | --- | --- | --- | --- | --- | --- | --- | --- |
| Rank based on IP | PI |  |  | PII |  |  | PIII |  |  |
|  | Feature | IP |  | Feature | IP |  | Feature | IP |  |
| 1 | Phase s.r. by MoA | 0.040 |  | Clinical trial cost | 0.047 |  | Company s.r. | 0.047 |  |
| 2 | Phase s.r. by indication | 0.038 |  | Phase s.r. by indication | 0.042 |  | Phase s.r. by indication | 0.047 |  |
| 3 | Company s.r. | 0.038 |  | Company s.r. | 0.042 |  | Phase s.r. by MoA | 0.045 |  |
| 4 | Clinical trial cost | 0.037 |  | Patent Duration | 0.040 |  | Clinical trial cost | 0.044 |  |
| 5 | Market s.r. | 0.036 |  | Market s.r. | 0.039 |  | Clinical trial results: Negative | 0.041 |  |
| 6 | Therapy type: Unclassified | 0.033 |  | Phase s.r. by MoA | 0.038 |  | Product Failed: No | 0.032 |  |
| 7 | R&D (Count) | 0.031 |  | Product Failed: Yes | 0.033 |  | Expedited status: Yes | 0.030 |  |
| 8 | Expedited status: No | 0.030 |  | Expedited status: Yes | 0.032 |  | Product Failed: Yes | 0.029 |  |
| 9 | Expedited status: Yes | 0.030 |  | Expedited status: No | 0.030 |  | Patent Duration | 0.029 |  |
| 10 | Product Failed: No | 0.026 |  | Product Failed: No | 0.028 |  | Expedited status: No | 0.027 |  |
| Feature importance by feature interactions |  |  |  |  |  |  |  |  |  |
| Rank based on VIC | PI |  |  | PII |  |  | PIII |  |  |
|  | Feature A | Feature B | VIC | Feature A | Feature B | VIC | Feature A | Feature B | VIC |
| 1 | Clinical trial cost | Phase s.r.by indication | 463 | Clinical trial cost | Company s.r. | 693 | Phase s.r.by indication | Company s.r. | 211 |
| 2 | Company Listed: Yes | Market s.r. | 408 | Company s.r. | R&D (Count) | 352 | Phase s.r.by MoA | Phase s.r.by indication | 202 |
| 3 | Therapy type: Unclassified | R&D (Count) | 280 | Patent duration | Company s.r. | 338 | Clinical trial results: Negative | Clinical trial cost | 174 |
| 4 | Therapy type: Monotherapy | R&D (Count) | 257 | Phase s.r.by MoA | Phase s.r.by MoA | 334 | Phase s.r.by MoA | Company s.r. | 161 |
| 5 | Phase s.r.by indication | Company s.r. | 248 | Clinical trial cost | Phase s.r.by MoA | 315 | Product Failed: No | Phase s.r.by MoA | 154 |
| 6 | Company s.r. | R&D (Count) | 230 | Phase s.r.by indication | Company s.r. | 297 | Clinical trial cost | Company s.r. | 137 |
| 7 | Clinical trial cost | Clinical trial cost | 226 | Company s.r. | Time in market | 285 | Company s.r. | R&D Cost | 130 |
| 8 | Phase s.r.by indication | R&D (Count) | 216 | Phase s.r.by MoA | Company s.r. | 281 | Product Failed: Yes | Company s.r. | 123 |
| 9 | Phase s.r.by MoA | Patents cite | 214 | Clinical trial cost | Company s.r. | 269 | Expedited status: Yes | Phase s.r.by MoA | 114 |
| 10 | Phase s.r.by MoA | Company s.r. | 211 | Time in Phase | Clinical trial cost | 259 | Expedited status: Yes | Phase s.r.by indication | 111 |

The Exhibit shows the top ranked features based on how frequently they were selected by BART. The BART tree inclusion proportion (IP) denotes the average fraction of times a feature was selected in a tree. The variable interaction count (VIC) sums how many times features were selected in consecutive nodes across BART trees. The higher the value of IP (VIC), the more relevant is the feature (feature combination) for classifying outcomes. For computational purposes it is the difference between the features' IPs and VICs, not their absolute values, that matters. The features are color coded based on their category: product features (orange), market features (blue), company features (pink), regulatory and other features (green). We abbreviate success ratio with s.r.

Having successfully trained and validated an ML algorithm, the next section will apply it to predict the outcome of the projects in the industry pipeline, i.e., those whose clinical research status was classified as on-going in our sample. To mitigate the potential bias from missing data we pre-processed the data using a nearest neighbor algorithm.

## 4. Predicting the outcome of compounds in the industry pipeline

Our project database contained 4,789 projects engaged in various phases of clinical research (PI = 1,858; PII = 2,372; PIII = 559) whose success or failure were not yet known. Exhibit 7 shows the predicted success rates for each phase and for all phases combined. Confidence intervals are shown by the black bars, whereas orange tics show the weighted average of the success rates derived from an analysis of the related literature based on observations between 2000 and 2018 (see Exhibit 24 in the supplementary material A.4).

**Exhibit 24:**
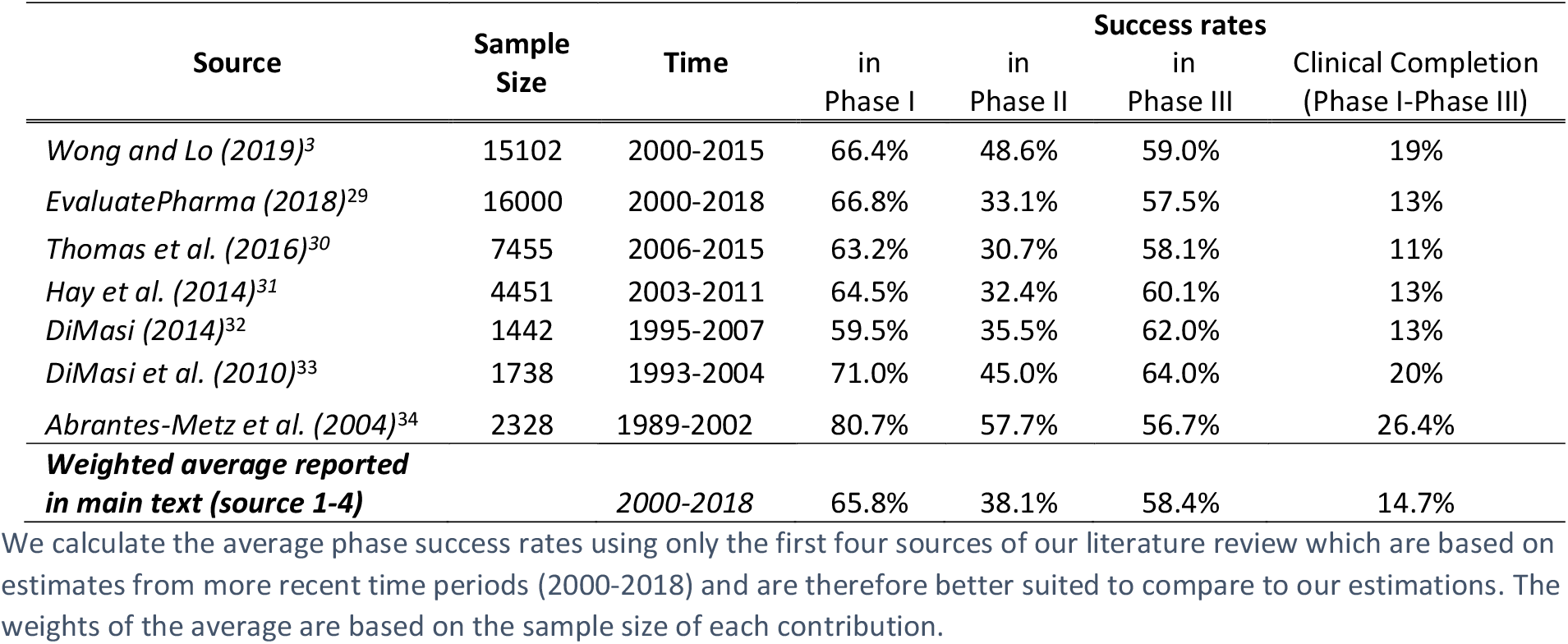
Analysis of the related literature regarding the estimation of phase success rates

Our predictions are slightly more optimistic than the average estimates of other independent research teams (clinical completion rates: 15.9% vs. 14.7%), in line with the increasing trend of drug approvals witnessed in recent years.^14^ Applying our project pipeline projections to a widely used R&D costing model^15^ suggests that the number of approvals in coming years will increase by 8.2% while the average cost of drug development will decrease by 3.9%.

This example illustrates an important point: the use of the algorithm does not by itself change project outcomes. It only predicts them with higher accuracy than existing methods. In this case, its predictions coincidentally agree with the estimates of other researchers. In addition, however, the algorithm provides detailed project information that allows R&D managers to reshape their pipelines to improve future success rates, which they can do by divesting projects with poor predicted outcomes and redirecting resources toward more promising ones.

For instance, Exhibit 8 shows that an orphan drug designation significantly boosts success rates over their expected average phase rates: +27 [+7] percentage points respectively for PII [PIII]; so do other expedited FDA review programs: +21% [+14%], MoA validated in a different indication: +17% [+13%]. If the molecule is already marketed in another indication, it boosts the success rate of a new indication by +22% [+22%] compared to expected average values. Lastly, projects associated with higher clinical trial expenses are estimated more likely to succeed +14% [+3%].^e^

Drug developers can also use the algorithm to study the impact of combinations of attributes and select the most desirable ones, or design better clinical research strategies. For example, the ML algorithm can be used to identify therapeutic areas and agents that offer better odds of clinical success. Exhibit 9 shows that these odds can vary considerably. In some instances small molecules or natural products seem to be less risky; in others, large molecule have the edge (see also Exhibit 25 and Exhibit 26 in the supplementary material A.4).

**Exhibit 9:** predicted phase II/III success rate per indication and technology

| Predicted success ratio | Phase II |  | Phase III |  |
| --- | --- | --- | --- | --- |
| Indication | Small molecules<br>Natural products | Large molecules | Small molecules<br>Natural products | Large molecules |
| Hepatic & biliary | 19.0% [58] | - [8] | - [6] | - [1] |
| Sensory organs | 22.7% [44] | 41.7% [36] | 35.7% [14] | - [7] |
| Cancer | 25.8% [690] | 30.8% [464] | 59.0% [83] | 64.8% [88] |
| Respiratory | 28.6% [35] | 33.3% [24] | - [4] | - [7] |
| Blood | 28.9% [38] | 53.3% [30] | 56.3% [16] | 63.6% [11] |
| Psychiatry | 36.8% [57] | - [0] | 33.3% [15] | - [0] |
| Immunology | 38.2% [34] | 33.3% [51] | 62.5% [16] | 60.0% [20] |
| Skin | 41.1% [73] | 45.8% [24] | 92.9% [14] | - [3] |
| Urinary tract | 42.1% [19] | - [10] | - [7] | - [3] |
| Diabetes | 43.8% [32] | 75.0% [12] | - [10] | - [3] |
| Neurology | 46.7% [120] | 35.7% [28] | 43.2% [44] | - [9] |
| Reproduction | 50.0% [18] | - | - [8] | - [1] |
| Musculoskeletal | 53.1% [32] | 44.7% [47] | 73.3% [15] | 66.7% [18] |
| Cardiovascular | 56.8% [37] | 45.2% [31] | 40.0% [20] | 41.7% [12] |
| Gastro-intestinal | 63.6% [44] | 37.5% [16] | - [10] | - [8] |
| Infections | 66.2% [77] | 32.4% [74] | 61.8% [34] | 45.5% [11] |
| HIV & related | - [8] | 28.6% [14] | - [6] | - [1] |
| Averages | 36.3% | 36.7% | 57.8% | 60.5% |
|  | 36.4% |  | 58.9% |  |

Number of observations in brackets. Success rates are not calculated for cells with fewer than 10 observations. Average success rates are weighted by the number of projects

**Exhibit 25:**
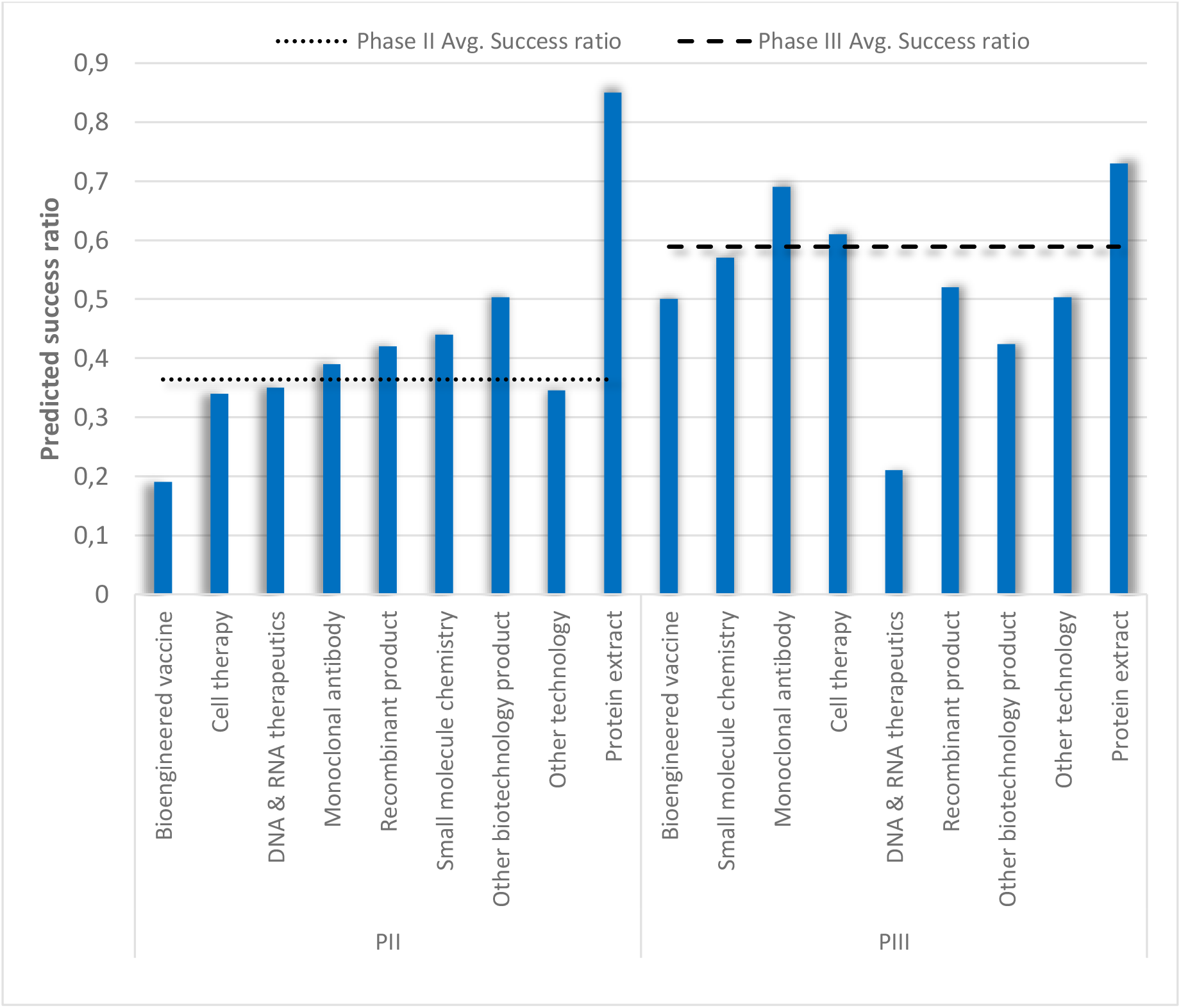
Predicted success rates of current PII/PIII pipeline by technology

**Exhibit 26:**
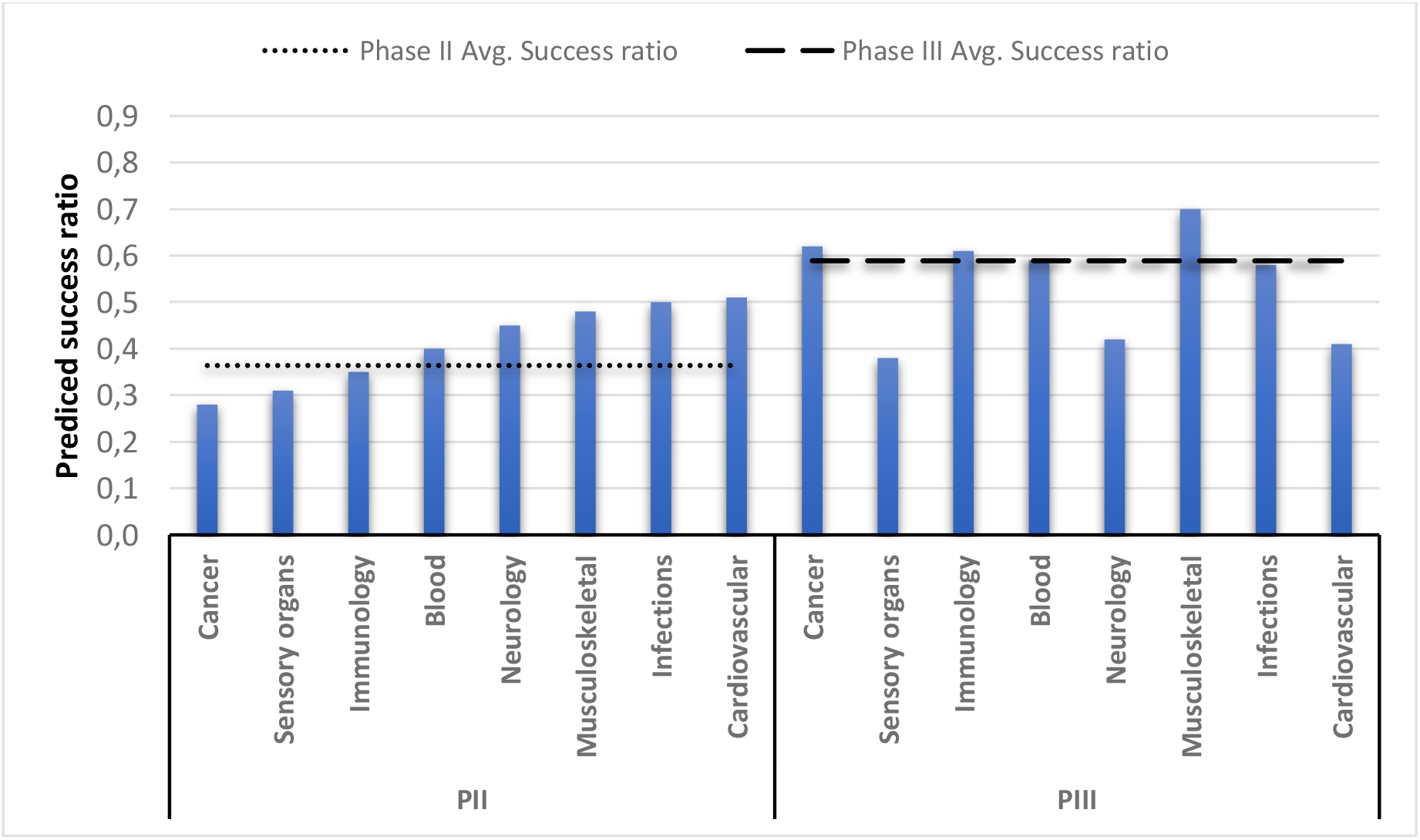
Predicted success rates of current PII/PIII pipeline by indication

## 5. Summary and discussion

We have evaluated the performance of different machine-learning algorithms to predict the clinical success (or failure) of individual pharmaceutical projects as they progress through the various phases of clinical research. The predictions of our “best-in-class” ML algorithm are substantially more accurate than traditional methods based on historic success rates or discriminant analysis. When predicting the outcome of pipeline projects, the average of our individual predictions accords with the aggregate historical benchmarks from the literature.

Our methodology closely adheres to good ML computational procedures, and additional steps were taken to control for the look-ahead bias, and filter out other potentially confounding factors such as drifts in the trends underpinning drug development, and the overweight influence of some indications. These robustness checks strengthen our findings and confirm the value of ML as a reliable predictor of clinical research outcomes^f^. Even though we abide by stringent quality standards, we should remain mindful that the algorithm may reproduce biases that can exist in the training set. Results should be inspected to detect such problem and corrective action be taken as appropriate. In other words, ML should inform the decision-making process of experts rather than replace it.

The implications of our work are important for individual companies, the pharmaceutical industry and the entire biomedical research ecosystem.

Pharmaceutical companies can use our approach to improve the quality of their pipelines by directing their R&D investment towards projects whose attributes makes them more likely to succeed. The algorithm’s capability to predict phase outcomes at the individual project level gives them a powerful tool to reorder their R&D priorities, and significantly boost their R&D productivity by raising new drug output and reducing the number and cost of failed trials.

At a higher level, our ML tool has the potential to change the industry’s risk profile. Over the decades, high risk has defined drug R&D. A handful of large companies has long dominated the industry because scale and staying power were required to survive high failure rates and the randomness of success. High prices and profitability were seen as necessary to withstand the devastating loss that a single failure could bring. Smaller companies found it difficult to develop enough new drugs to grow and rival their larger competitors. The ML demonstrated in this paper has the potential to change this. If risk is lower, more companies, especially smaller ones are likely to engage in drug R&D. If failures are fewer, less capital will be needed to succeed. That will stimulate entrepreneurial activity and cause the locus of innovation to gradually migrate from a handful of large companies to many smaller ones. The composition of innovation could also be affected since smaller, nimbler companies are more prone to explore new biology where high-value innovation has often been found. Scale and the resulting costs, risk-aversion, and bureaucracy could put large companies at a competitive disadvantage. The rationale for high prices will weaken and could evaporate. The result could be an industry that is more entrepreneurial, more productive, and cheaper.

ML could also bring significant changes to the broader biomedical research ecosystem. It could divert resources away from some diseases and therapeutic areas that do not have the attributes they need to score well with the algorithm. This could happen, for instance, if the drug’s mode of action has not been validated, which is often the result of a poorly understood pathology. It would be a signal to policymakers and academic researchers to reorder their priorities and increase funding and focus on those areas where innovation is unlikely to flourish until knowledge gaps have been filled. The ecosystem will become smarter. It will have a tool to allocate resources where they are most needed in both basic and translational research.

Lastly, this paper is another successful step in using ML to address challenges that, until now, have often been seen as intractable. BART had previously been used to address such challenges – for instance, the prediction of movie box-office revenues^16^. Here we apply it to predict the successes and failures in clinical development, a problem that long vexed the pharmaceutical industry, despite its capabilities and the obvious economic value of such tool. These successes raise hopes that further ML-driven breakthroughs are at hand. However, achieving them will require access to vast amounts of high-quality data to train algorithms – both positive data about successful experiments as well as negative data about failures. Assembling them will require extensive data-sharing. Still, despite well-intended policies at funding organizations, there is concern that data-sharing at many organizations remains half-hearted^17, 18^. To reap the full societal benefits of ML, this needs to change.

## Data Availability

Data is proprietary and cannot be shared publicly

## Supplementary information A

### 1. Data set characteristics

### 2. Machine learning routine and performance evaluation

#### Machine learning methods used

The fate of a pharmaceutical project as it passes through the various phases of clinical research depends on a combination of product-, company-, and market attributes. These attributes can be used by machine learning algorithms to predict the most likely outcome. Since the performance of machine learning algorithms is highly data-dependent, we train eight different algorithms using training data from three sets of projects belonging to phase I, II, and III. We then evaluate the performance of the trained algorithms by analyzing their ability to discriminate between successful and failed projects when applied to three validation sets for phase I, II, and III.

The eight ML algorithms span methods that are frequently used for prediction tasks, i.e., a simple decision tree (DT), boosted decision trees (C5.0)^19^, a random forest algorithm (RF)^20^, a Bayesian additive regression tree (BART)^10,21^, a support vector machine (SVM)^22^, an artificial neural network (ANN)^23^ a linear probabilistic regression (PROBIT), and an ensemble learner based on the three best performing methods.

DT, C5.0, RF and BART are tree-based classification methods, which are suited to problems where non-linearities and interactions between features are plausible, but unknown. A classification tree can be thought of as a set of successive decision rules, called nodes. The branches, that extend from the nodes, split the observations according to these decision rules. At the terminal nodes each observation is categorized as either success or failure.

The DT algorithm relies on only one tree while C5.0, RF and BART create an ensemble of trees but in different ways. The C5.0 method uses gradient boosting that enables the algorithm to learn from classification errors of prior trees; RF averages across estimates from multiple trees based on a random subset of features and projects; and BART sums the contribution of multiple trees. The structure of these trees depends on Bayesian priors that, to prevent overfitting of the model, are also applied on the error variance. The tree regularization achieved by the Bayesian approach combined with limiting the sum of trees acts as a natural way to prevent features from entering the model that add little explanatory power (i.e., in case of multicollinearity). Moreover, BART incorporates a Missingness-Incorporated-in-Attributes procedure (MIA)^12^, which expands the predictor space to include information on missing features (we elaborate on this below).

The SVM algorithm, on the other hand, classifies observations by fitting a hyperplane to the dataset that divides it into predicted successes and failures. The hyperplane is supported by vectors which are chosen so that the overall distance (called the margin) between the hyperplane and the two classes is maximized along with the prediction accuracy.

The ANN algorithm operates by constructing a network of nodes (called neurons), which are autonomous data-processing units. The neurons are organized into three or more layers: an input layer that receives the input data, one or more downstream hidden layers, and an output layer that produces the predicted values. Each neuron receives incoming signals, processes them, and sends outgoing signals to other neurons. Each neuron processes incoming signals by using an activation function that resembles that of biological neurons, i.e., if the signal is below a threshold, it is not transmitted; if it is above, it is modulated according to a function that is characteristic of each neuron, and passed forward. During the iterative training process, the network receives pairs of actual input and output data. The input data is converted into signals by the neurons of the input layer. Those signals are sent to other neurons in downstream hidden layers which reprocess them and send them onward until they reach the output layer, where they are converted into output values. At each iteration, the modulation of each neuron’s signal is adjusted in a way that lessens the distance between the predicted and actual output value(s), thereby improving the quality of the prediction, until the training is completed.

Next,, we train a standard linear regression PROBIT model to compare how classification performance changes, when outcomes are predicted by a linear combination of features without allowing for variable interactions. Lastly, we construct an ensemble learner based on a weighted average of the predictions of the three best performing methods. The weights are chosen via cross validation on part of the training set such that the linear combination of single predictions minimizes the prediction error.

#### Machine learning training procedure

Each of the three datasets is randomly split into a training set (70%) used to train the algorithms and a validation set (30%) used afterward to assess the performance of the trained algorithms. The validation set is not used until the training has been completed. Exhibit 12 sketches the training and validation procedure used in this paper.

Before training the algorithms, we analyze whether pre-processing the data improves predictive performance on the training set. Since missing data can negatively affect the performance of the trained algorithm, it is important to examine the volume of missing data and ways to mitigate it (see Exhibit 11 for a visual presentation of missingness across data sets). Applying Little’s tests to the feature space across data sets, allows us to clearly reject the Null hypothesis that missing data are randomly distributed^24^. There are many ways to handle missing data in input datasets. Here, we consider three approaches: a complete case (CC) analysis, in which features with more than 70% of missing values and all remaining observations that contain missing values are excluded, a nearest neighbor (NN) imputation algorithm, and an internal imputation (II) using directly the respective algorithm (not available for PROBIT, SVM and ANN). We set the number of neighbor observations considered in NN to 5 (5NN), since 5NN combined with RF achieves good classification performance in a similar setting^5^. Each training set is randomly split into a training set (70%) on which each algorithm is trained while successively applying each of the three missing value handling techniques. The trained algorithms are then evaluated on the remaining 30% of the training data by calculating the area under the receiver operating characteristic curve (AUC). This helps select the missing value handling technique that produces the highest AUC for each algorithm without involving the original validation dataset. In this case, the highest AUC is achieved by 5NN imputation for C5.0, PROBIT, SVM and ANN while the other algorithms are more accurate when using the internal missing value routines embedded in their software (see Exhibit 13). Missing input data values are imputed without considering their impact on the success classification, to prevent the imputed values from somehow reflecting this information. Training and validation datasets are imputed separately to avoid inducing any form of relation between them.

**Exhibit 13:**
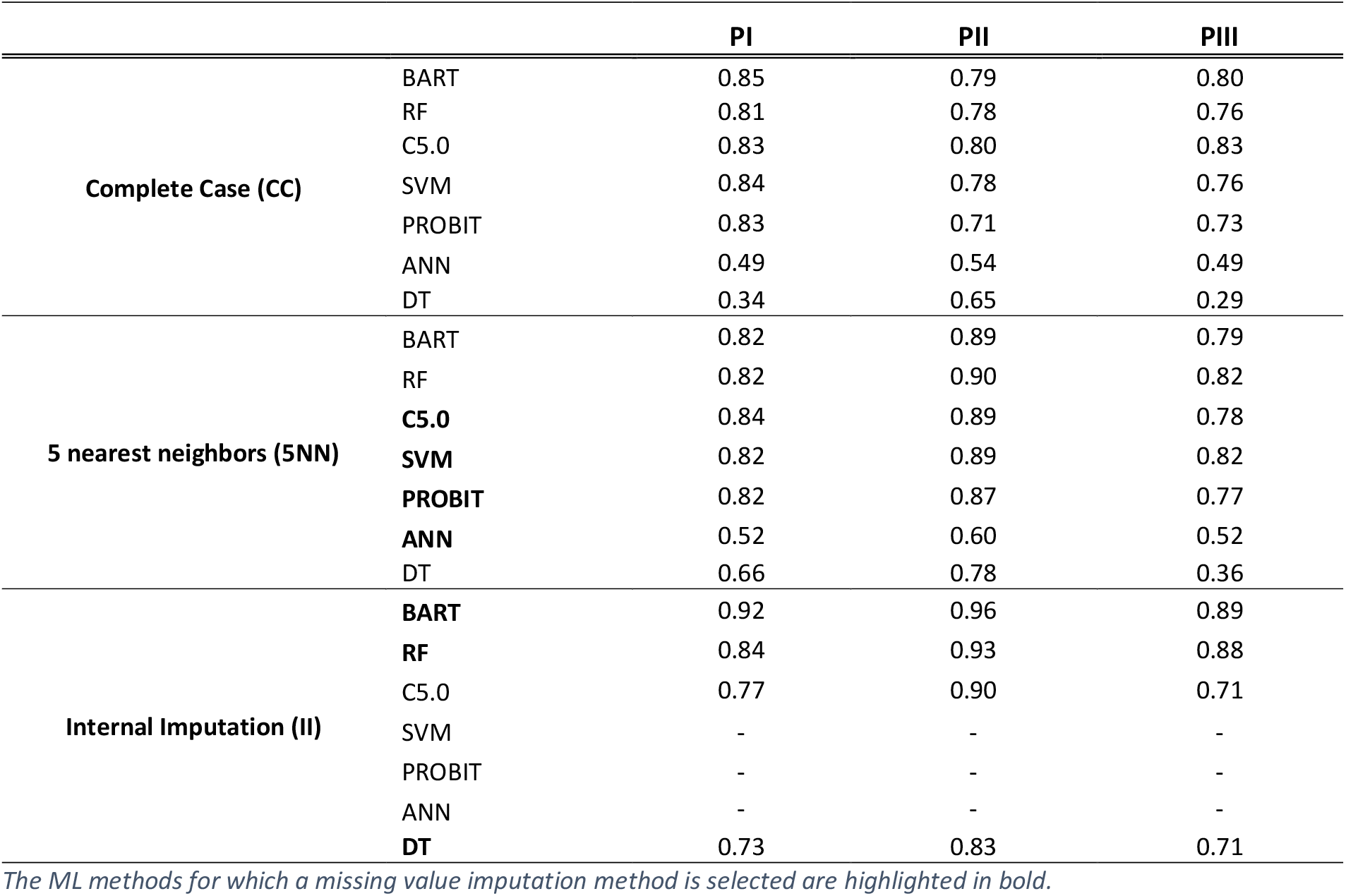
Missing value imputation techniques by AUC across ML methods and data sets

After imputing missing values, we perform a feature selection step to keep excessive feature inclusion from degrading the predictive performance of the algorithms (e.g. by avoiding multicollinearity in selected features). We evaluate three feature selection methods: LASSO^25^, an oft-applied method that is based on the shrinkage of linear regression coefficients, RF_SE, an iterative variable elimination method used in RF and based on the smallest prediction loss^26^ and BART_IP that selects variables based on their inclusion proportion in a BART algorithm with a small number of trees^13^. After completing the imputation of the missing values, we use 70% of the training data to train each algorithm on each data set for each feature selection technique and compare the results based on the AUC from the 30% remaining training data (see Exhibit 14). For PROBIT, DT and ANN the RF_SE method performs best whereas LASSO is selected for SVM. The ensemble tree methods BART, RF and C5.0 perform best without the use of an additional feature selection step.

**Exhibit 14:** Feature selection techniques by AUC across ML methods and data sets

*The ML methods for which a missing value imputation method is selected are highlighted in bold.* Exhibit 14: Feature selection techniques by AUC across ML methods and data sets
| Feature selection technique | ML Method | PI | PII | PIII |
| --- | --- | --- | --- | --- |
| LASSO | BART | 0.90 | 0.96 | 0.90 |
|  | RF | 0.84 | 0.93 | 0.87 |
|  | C5.0 | 0.83 | 0.89 | 0.82 |
|  | **SVM** | 0.83 | 0.89 | 0.83 |
|  | PROBIT | 0.83 | 0.88 | 0.79 |
|  | ANN | 0.82 | 0.85 | 0.76 |
|  | DT | 0.74 | 0.84 | 0.71 |
| RF_SE | BART | 0.89 | 0.94 | 0.89 |
|  | RF | 0.84 | 0.92 | 0.86 |
|  | C5.0 | 0.81 | 0.88 | 0.82 |
|  | SVM | 0.82 | 0.89 | 0.83 |
|  | **PROBIT** | 0.83 | 0.88 | 0.84 |
|  | **ANN** | 0.82 | 0.88 | 0.50 |
|  | **DT** | 0.73 | 0.85 | 0.71 |
| BART_IP | BART | 0.90 | 0.95 | 0.91 |
|  | RF | 0.85 | 0.92 | 0.87 |
|  | C5.0 | 0.75 | 0.88 | 0.81 |
|  | SVM | 0.79 | 0.88 | 0.80 |
|  | PROBIT | 0.78 | 0.88 | 0.81 |
|  | ANN | 0.76 | 0.87 | 0.78 |
|  | DT | 0.73 | 0.84 | 0.71 |
| No Feature selection | **BART** | 0.92 | 0.96 | 0.89 |
|  | **RF** | 0.84 | 0.93 | 0.88 |
|  | **C5.0** | 0.84 | 0.89 | 0.78 |
|  | SVM | 0.82 | 0.89 | 0.82 |
|  | PROBIT | 0.82 | 0.87 | 0.77 |
|  | ANN | 0.52 | 0.60 | 0.52 |
|  | DT | 0.73 | 0.83 | 0.71 |

The ML methods for which a feature selection technique is selected are highlighted in bold.

In a last step before the final validation, some algorithm hyperparameters are tuned using 5fold-cross validation on the complete training data for each data set. Hyperparameters tuned are the number of trees and the cut-off probability in BART, the number of randomly sampled variables at each split in RF, the number of boosting iterations in C5.0, the kernel shape of the distance measure in SVM and the number of hidden layers in ANN. We report the tuning results in Exhibit 15 to show that performance characteristics do not vary substantially across evaluated hyperparameter ranges. For each algorithm and data set we choose the hyperparameter specification that performed best.

**Exhibit 15:** Average 5-fold cross validation results for tuning hyper parameters across algorithms

The ML methods for which a feature selection technique is selected are highlighted in bold. Exhibit 15 Average 5-fold cross validation results for tuning hyper parameters across algorithms
|  | PI | PII | PIII |
| --- | --- | --- | --- |
| **Cutoff- prob rule class** | **BART (Accuracy)** |  |  |
| 0.3 | 0.829 | 0.881 | 0.856 |
| 0.4 | 0.840 | 0.892 | **0.892** |
| 0.5 | **0.848** | **0.903** | 0.879 |
| 0.6 | 0.840 | 0.892 | 0.851 |
| 0.7 | 0.817 | 0.867 | 0.792 |
| **# trees** |  |  |  |
| 50 | 0.850 | 0.899 | 0.875 |
| 100 | 0.855 | **0.901** | **0.877** |
| 150 | **0.859** | 0.900 | 0.870 |
| 200 | 0.856 | 0.897 | 0.870 |
| **# sampled variables** | **RF (Accuracy)** |  |  |
| 5 | 0.840 | 0.879 | 0.831 |
| 6 | 0.835 | **0.879** | **0.838** |
| 7 | 0.837 | 0.876 | 0.834 |
| 8 | **0.841** | 0.876 | 0.824 |
| **Boosting iterations** | **C5.0 (Accuracy)** |  |  |
| 10 | 0.793 | 0.847 | 0.795 |
| 20 | 0.804 | 0.846 | 0.795 |
| 30 | 0.807 | 0.850 | 0.806 |
| 40 | 0.811 | 0.851 | 0.811 |
| 50 | 0.811 | 0.852 | **0.813** |
| 60 | 0.810 | 0.849 | 0.810 |
| 70 | 0.812 | 0.850 | 0.800 |
| 80 | 0.812 | 0.852 | 0.803 |
| 90 | **0.814** | 0.851 | 0.803 |
| 100 | 0.812 | **0.853** | 0.805 |
| **Kernel shape** | **SVM (AUC)** |  |  |
| linear | 0.849 | 0.888 | 0.880 |
| polynomial | 0.837 | 0.888 | 0.878 |
| radial | **0.852** | **0.899** | **0.892** |
| sigmoid | 0.839 | 0.892 | 0.886 |
| **# hidden layers** | **ANN (AUC)** |  |  |
| 1 | 0.528 | 0.878 | **0.865** |
| 2 | **0.720** | 0.874 | 0.857 |
| 3 | 0.640 | 0.854 | 0.856 |
| 4 | 0.718 | **0.878** | 0.856 |
| 5 | 0.552 | 0.867 | 0.846 |
| 6 | 0.691 | 0.867 | 0.844 |
| 7 | 0.686 | 0.873 | 0.833 |
| 8 | 0.665 | 0.878 | 0.809 |
| 9 | 0.680 | 0.837 | 0.837 |
| 10 | 0.573 | 0.859 | 0.834 |

The hyperparameters that correspond to the highest performance for each phase (highlighted in bold) are chosen in the final training of the algorithms.

Every algorithm is then trained on the full training dataset using the best performing missing value technique, feature selection criterion and adjusted hyperparameters. The trained algorithms are subsequently assessed using the validation data set that has been kept separate from the training procedure. To rule out that the test results are influenced by the random selection of the validation data, the entire training and validation procedure is repeated 100 times for each model and each data set and average performance measures are reported.

#### Machine learning performance validation

For all algorithms and data sets we report multiple performance features. The AUC measure is frequently used to report classification performance since its value is independent of the choice of a specific classification threshold.^9^ We report the average AUC together with the mean 95% confidence interval, calculated by the Delong method for each repetition individually and then averaged over all obtained upper and lower bounds.^11^ The higher the AUC, the better the algorithm solves the trade-off between type I and type II prediction errors. An AUC of 0.5 indicates that a random outcome assignment would be equally predictive. An AUC of 1 indicates that there exists at least one classification threshold at which the model classifies each case correctly. Besides the AUC, the algorithm’s sensitivity (SENS, the number of correctly classified successes over the total number of true successes) and its specificity (SPEC, the number of correctly classified failures over the total number of true failures), the positive predictive value (PPV, the number of correctly classified successes over the total number of classified successes), the negative predictive value (NPV, the number of correctly classified failures over the total number of classified failures), the F1 score (F1, the harmonic mean between PPV and SENS), and the balanced accuracy (BACC, the geometric mean between SENS and SPEC) are reported in Exhibit 16. SENS, SPEC, PPV and NPV are intuitive performance measures stemming directly from the confusion matrix. The last two measures are useful to analyze in case the data is unbalanced in terms of outcomes.

In all three data sets the BART algorithm achieves the highest classification performance with an average AUC of 0.93 in PI, 0.96 in PII, and 0.94 in PIII. It also performs best in terms of F1 (0.90 PI, 0.86 PII, and 0.90 PIII) and BACC (0.83 PI, 0.89 PII, and 0.86 PIII) We therefore refer to this algorithm as ‘best-in-class’ in the main text and use it during subsequent analysis. RF shows a lower performance than BART for most measures apart from Sensitivity and NPV in PI and PIII. Neither algorithm relies on additional feature selection or missing value imputation techniques, which makes them powerful stand-alone tools in our analysis. For C5.0 we find a slightly lower average AUC than for RF, followed by SVM. The performance of PROBIT is similar to SVM which shows that careful choice of missing value imputation and feature selection techniques can offset the linear restrictions imposed by this model. The worst performing methods in terms of average AUC and BACC are ANN and DT, presumably because in our application they were overfitting the data during the training phase. Lastly, the ensemble learner, constructed from optimally weighting the prediction values of BART, RF and C5.0 on the training set (Exhibit 17 reports the weights), performs worse than the single BART algorithm in terms of AUC in PI and PII. Looking additionally at balanced performance measures such as F1 and BACC, the stand-alone BART method performs better across phases, which is why we selected it for the main part of our analysis.

**Exhibit 17:**
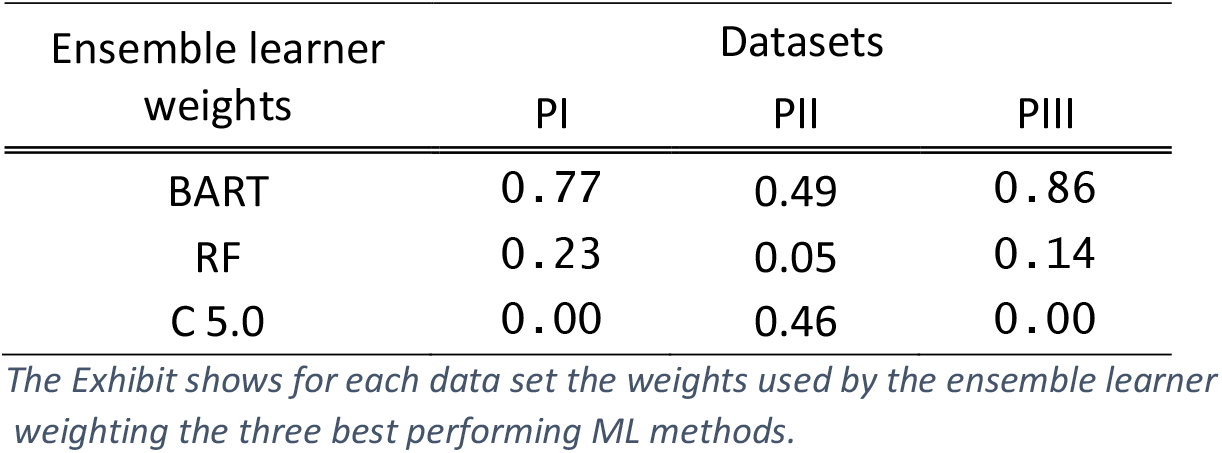
Weights of ensemble learner across data sets

#### Discriminant analysis for feature selection and classification

In practice, classification problems are often approached using methods whose input parameters relate linearly to outcomes, which we broadly refer to as discriminant analysis (DISCR). As one specific example of such a linear discriminant analysis, we implement a backward/forward probabilistic regression procedure with Bayesian information criterion and compare its classification performance on the validation set with that of BART in the main text.

Bayesian information criterion (BIC) evaluates how well a model explains the data while staying as parsimonious as possible. The better this tradeoff gets solved by a model the higher is its BIC value. The procedure optimizes the BIC value by adding or removing features to a probabilistic regression, which means it finds the model that explains the data best without including too many parameters. The procedure stops when neither adding nor removing features contributes to the BIC of the model, thus keeping only the features with the highest explanatory power.

We report the coefficients of the selected features, their standard errors and p-values in Exhibit 18, which complements the feature importance measures elicited by BART, because directionality and significance of effects are easily interpreted. Yet, one needs to bear in mind that the model is assumed to be linear in the effect of features on project outcome which might not be appropriate given its moderate predictive performance.

**Exhibit 18:**
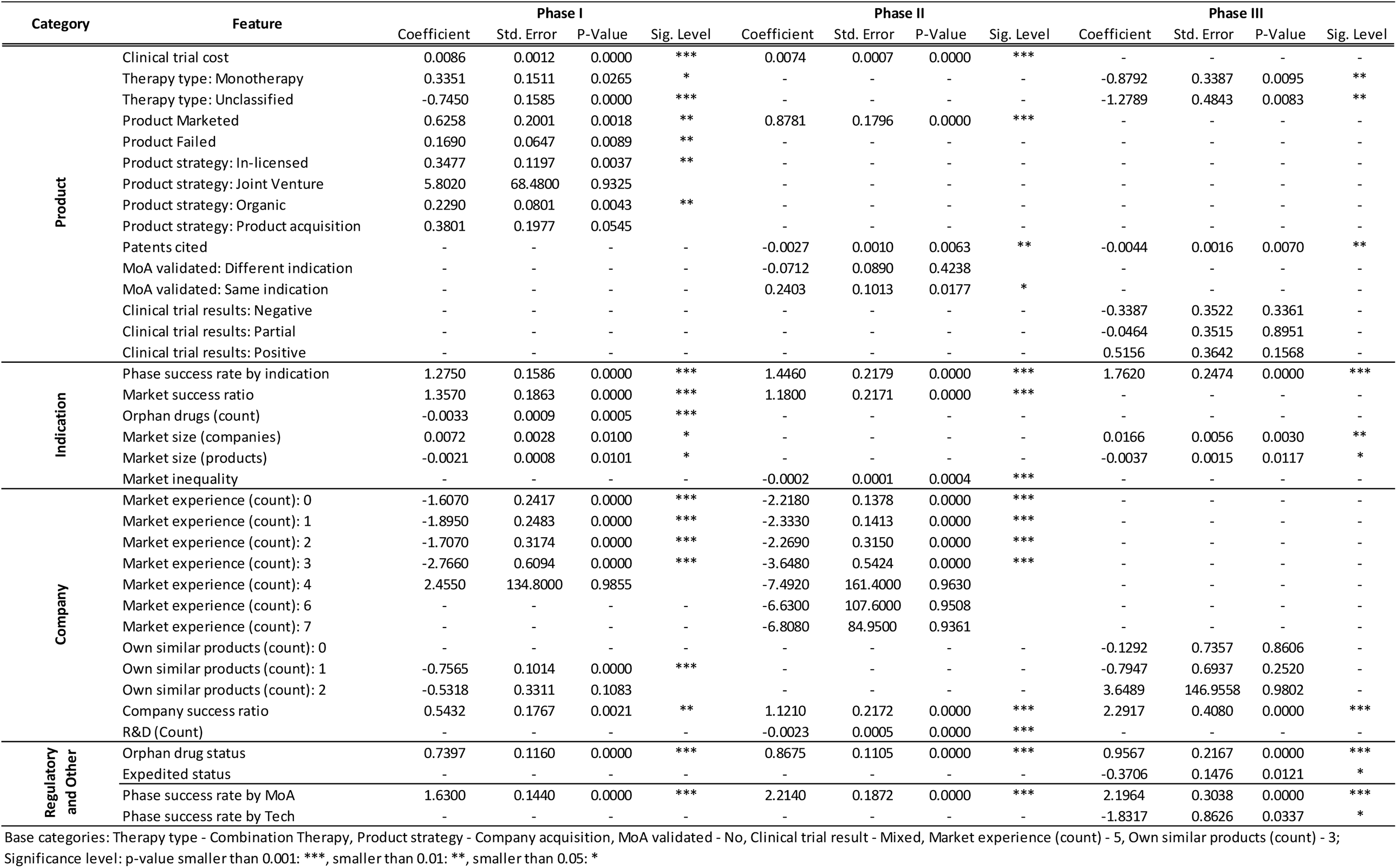
Most important features based on backward/forward probabilistic regression

During the validation task, the coefficients derived from running the DISCR model on the training set are applied to the input data of the validation set, resulting in classification values that can be compared to the true outcomes via various performance measures (see Exhibit 4 in main text) or classification plots (Exhibit 23 in the supplementary material A.4).

#### Machine Learning for predicting project outcomes of the current pipeline

When predicting project outcomes of the current pipeline, one cannot directly validate the resulting predictions. Therefore, it is crucial that the training set and the current pipeline share the same properties with respect to missing values and evaluated features. For on-going projects, feature information is on average better than for historical projects (even though information on some features such as clinical trial results is not fully available – see Exhibit 11), since for current projects an open data approach has been enforced.^27^ To rule out that the difference of missingness in the data influences prediction results, we first impute labeled and unlabeled data separately using a 5NN algorithm (see missing value imputation techniques above). We then train the BART algorithm and analyze the out-of-sample performance on the separately kept validation set (performance according to BACC: PI=76%, PII=81%, PIII=78%). Since the BART algorithm performs optimally when it imputes missing values internally, using 5NN to ensure that missingness between training and prediction data is comparable results in a slightly lower yet still advantageous classification performance compared to other methods. Next, the algorithm gets trained on the complete set of labeled data (see above) and is eventually used to predict outcomes of on-going projects. The results of this process are depicted in Exhibit 7, Exhibit 8, and Exhibit 9 of the main text and Exhibit 25, Exhibit 26 in the supplementary material A.4.

### 3. Result robustness checks

Even though our experimental set-up closely follows common ML procedures and the obtained project classification on validation sets seem very promising, we need to rule out that the reported results are driven by particularities in the data or the training/validation routine. We therefore perform various changes in the training and validation approach that aim at providing insights into the robustness of our validation results.

#### Time series validation technique

Randomly sampling training and validation sets from the data could lead to so called look-ahead bias, meaning that algorithms are trained on projects that happened later in time and thus learn from future information. To mitigate look-ahead bias we perform an additional time-series training and testing routine^28^ and compare its performance to our base results.

To implement the time series validation technique, we train the algorithms on all projects whose status has been determined prior to year *t* and validate the performance using projects only in year *t*. Making sure to have enough observations in every training and testing set, the algorithms are first trained on all projects with determined outcome between 2009 and 2014, and then validated using projects whose outcome determined 2015 (validation set is referred to as 2015). For the second (third) window, the algorithms are trained on projects determined between 2009 and 2015 (2009 and 2016) and validated on projects determined in 2016 (2017), and so on.

We opt for this time series approach using multiple training and validation sets, to observe how the prediction quality of the ML methods shift over time. We compare its prediction performance with the performance obtained from our base results (randomly splitting of data 100 times) using BACC (see Exhibit 19) and AUC (see Exhibit 20) on all algorithms except the ensemble learner (we feared that the data quantity would not be sufficient to learn the optimal weights on a separate training set). We find that the time series approach performs worse in PI, than the random sampling, which allows the conclusion that look-ahead bias is at least partly responsible for the classification results in PI. This is not surprising, since information on PI clinical trials are voluntarily disclosed which may induce some lags in information reporting of failed trials that are picked up by the algorithms using random sampling. In PII and PIII data sets, we do not detect signs of look-ahead bias when comparing the random sampling performance to the time series approach (AUC of BART in PI under random sampling: 93% vs. under mean AUC time series: 91%; PII 96% vs. 96% and PIII: 94% vs. 98%), i.e. splitting training and testing data according to a time dimension does not deliver worse results suggesting that look-ahead bias in PII and PIII data is less of a concern.

**Exhibit 19:** Time series validation technique - algorithm performance according to balanced accuracy

| PI |  |  |  |  |  |  |
| --- | --- | --- | --- | --- | --- | --- |
|  | 2015 | 2016 | 2017 | 2018 | Mean | BACC by<br>Random Split |
| BART | 0.750 | 0.731 | 0.622 | 0.777 | **0.720** | **0.835** |
| RF | 0.752 | 0.709 | 0.587 | 0.604 | 0.663 | 0.751 |
| C 5.0 | 0.779 | 0.760 | 0.638 | 0.687 | 0.716 | 0.750 |
| SVM | 0.705 | 0.743 | 0.607 | 0.666 | 0.680 | 0.725 |
| PROBIT | 0.749 | 0.674 | 0.576 | 0.850 | 0.712 | 0.730 |
| ANN | 0.752 | 0.688 | 0.583 | 0.819 | 0.710 | 0.708 |
| DT | 0.737 | 0.706 | 0.554 | 0.850 | 0.712 | 0.677 |
| PII |  |  |  |  |  |  |
|  | 2015 | 2016 | 2017 | 2018 | Mean | BACC by<br>Random Split |
| BART | 0.852 | 0.932 | 0.903 | 0.859 | **0.887** | **0.886** |
| RF | 0.685 | 0.756 | 0.877 | 0.741 | 0.765 | 0.853 |
| C 5.0 | 0.784 | 0.887 | 0.867 | 0.819 | 0.839 | 0.823 |
| SVM | 0.755 | 0.892 | 0.850 | 0.809 | 0.827 | 0.799 |
| PROBIT | 0.848 | 0.893 | 0.851 | 0.857 | 0.862 | 0.790 |
| ANN | 0.859 | 0.910 | 0.830 | 0.847 | 0.861 | 0.716 |
| DT | 0.789 | 0.711 | 0.764 | 0.853 | 0.779 | 0.751 |
| PIII |  |  |  |  |  |  |
|  | 2015 | 2016 | 2017-2018 |  | Mean | BACC by<br>Random Split |
| BART | 0.786 | 0.798 | 0.931 |  | **0.838** | **0.862** |
| RF | 0.786 | 0.688 | 0.810 |  | 0.761 | 0.772 |
| C 5.0 | 0.685 | 0.688 | 0.883 |  | 0.752 | 0.778 |
| SVM | 0.628 | 0.594 | 0.905 |  | 0.709 | 0.759 |
| PROBIT | 0.670 | 0.781 | 0.823 |  | 0.758 | 0.713 |
| ANN | 0.670 | 0.704 | 0.918 |  | 0.764 | 0.734 |
| DT | 0.798 | 0.721 | 0.858 |  | 0.793 | 0.727 |

The table shows the BACC across algorithms using as validation set the projects of the year in the respective column and as training set the projects whose outcome was determined prior to that year. "Mean" denotes the average BACC across yearly testing sets. "BACC by Random Split" corresponds to the last column of Exhibit 16 to facilitate comparison.

**Exhibit 20:** Time series validation technique - algorithm performance according to AUC

| PI |  |  |  |  |  |  |
| --- | --- | --- | --- | --- | --- | --- |
|  | 2015 | 2016 | 2017 | 2018 | Mean | AUC by<br>Random Split |
| BART | 0.945 | 0.919 | 0.854 | 0.925 | 0.911 | 0.934 |
| RF | 0.890 | 0.850 | 0.741 | 0.885 | 0.841 | 0.883 |
| C 5.0 | 0.936 | 0.906 | 0.806 | 0.851 | 0.875 | 0.864 |
| SVM | 0.907 | 0.871 | 0.783 | 0.888 | 0.862 | 0.855 |
| PROBIT | 0.850 | 0.829 | 0.674 | 0.883 | 0.809 | 0.843 |
| ANN | 0.844 | 0.829 | 0.735 | 0.887 | 0.824 | 0.752 |
| DT | 0.807 | 0.719 | 0.457 | 0.833 | 0.704 | 0.693 |

| PII |  |  |  |  |  |  |
| --- | --- | --- | --- | --- | --- | --- |
|  | 2015 | 2016 | 2017 | 2018 | Mean | AUC by<br>Random Split |
| BART | 0.965 | 0.977 | 0.965 | 0.949 | 0.964 | 0.960 |
| RF | 0.913 | 0.942 | 0.948 | 0.825 | 0.907 | 0.926 |
| C 5.0 | 0.932 | 0.955 | 0.955 | 0.927 | 0.942 | 0.913 |
| SVM | 0.927 | 0.972 | 0.950 | 0.933 | 0.945 | 0.897 |
| PROBIT | 0.928 | 0.958 | 0.936 | 0.925 | 0.936 | 0.886 |
| ANN | 0.924 | 0.970 | 0.890 | 0.891 | 0.919 | 0.770 |
| DT | 0.900 | 0.859 | 0.457 | 0.833 | 0.762 | 0.850 |

| PIII |  |  |  |  |  |  |
| --- | --- | --- | --- | --- | --- | --- |
|  | 2015 | 2016 | 2017-2018 |  | Mean | AUC by<br>Random Split |
| BART | 0.971 | 0.977 | 0.978 |  | 0.975 | 0.944 |
| RF | 0.937 | 0.901 | 0.944 |  | 0.927 | 0.908 |
| C 5.0 | 0.941 | 0.925 | 0.950 |  | 0.939 | 0.882 |
| SVM | 0.924 | 0.918 | 0.983 |  | 0.942 | 0.866 |
| PROBIT | 0.870 | 0.964 | 0.930 |  | 0.921 | 0.831 |
| ANN | 0.916 | 0.929 | 0.970 |  | 0.938 | 0.821 |
| DT | 0.840 | 0.870 | 0.927 |  | 0.879 | 0.791 |

The table shows the AUC across algorithms using as validation set the projects of the year in the respective column and as training set the projects whose outcome was determined prior to that year. "Mean" denotes the average AUC across yearly testing sets. "AUC by Random Split" corresponds to the last column of Exhibit 16 to facilitate comparison.

#### Performance on recent data only

Since the drug development landscape changes over time, it might be useful to restrict the training/validation observations to the most recent projects. That way we guarantee that the algorithms are not trained on data, which contains information that might be outdated. Moreover, using only the most recent observations we avoid issues that relate to static project features (features that reflect information at the time the data was sourced rather than when the project outcome was determined – see Exhibit 10 for a description of features). This is a common problem when working with historical information dating back a few years, since much information was simply not digitalized and is now not possible to obtain ex-post.

We train and validate each algorithm using data only on the two latest years limiting the issues discussed above while guaranteeing enough observations for the training and testing routine. The algorithms are used as specified previously (see section: in supplementary material A.2: Machine learning training procedure) and validation results are sampled 100 times randomly splitting the training/testing data at each run. We run the robustness check for each algorithm except the ensemble learner which would require additional training data to optimally select the weights of algorithm predictions. The performance of ML techniques on the reduced dataset using only recent data (see Exhibit 21) looks similar to the ones in Exhibit 16 (validation results using all data). BART is still the best performing methodology to be used for projects in phases II and III. As for phase I, C 5.0 performs best in terms of BACC but not AUC. All in all, we do not find any major differences in our results due to historical observations in our results and we confirm BART as our ‘best-in-class’ approach.

**Exhibit 21:** Average validation results across ML algorithms and data sets – using only data of the two most recent years

| ML Method | Data set | AUC Mean | AUCL Mean | AUCH Mean | SENS Mean | SPEC Mean | PPV Mean | NPV Mean | F1 Mean | BACC Mean |
| --- | --- | --- | --- | --- | --- | --- | --- | --- | --- | --- |
| BART | PI | **0.90** | **0.83** | **0.97** | 0.99 | 0.30 | 0.90 | 0.89 | 0.94 | 0.65 |
|  | PII | **0.96** | **0.91** | **1.00** | 0.95 | 0.76 | **0.89** | **0.88** | **0.92** | **0.85** |
|  | PIII | **0.99** | **0.97** | **1.00** | 0.97 | **0.90** | **0.95** | 0.94 | **0.96** | **0.94** |
| RF | PI | 0.84 | 0.73 | 0.94 | 0.99 | 0.35 | 0.90 | 0.85 | **0.94** | 0.67 |
|  | PII | 0.91 | 0.84 | 0.98 | **0.96** | 0.58 | 0.83 | **0.89** | 0.89 | 0.77 |
|  | PIII | 0.97 | 0.91 | **1.00** | 0.97 | 0.68 | 0.86 | 0.92 | 0.91 | 0.82 |
| C 5.0 | PI | 0.85 | 0.76 | 0.94 | 0.97 | **0.43** | **0.91** | 0.73 | 0.94 | **0.70** |
|  | PII | 0.92 | 0.86 | 0.99 | 0.92 | 0.74 | 0.88 | 0.83 | 0.90 | 0.83 |
|  | PIII | 0.94 | 0.86 | **1.00** | 0.94 | 0.81 | 0.92 | 0.90 | 0.93 | 0.88 |
| SVM | PI | 0.81 | 0.71 | 0.92 | **1.00** | 0.20 | 0.88 | **0.91** | 0.94 | 0.60 |
|  | PII | 0.94 | 0.88 | 0.99 | 0.92 | **0.76** | 0.89 | 0.83 | 0.90 | 0.84 |
|  | PIII | 0.98 | 0.93 | **1.00** | **0.99** | 0.80 | 0.91 | **0.98** | 0.95 | 0.89 |
| PROBIT | PI | 0.74 | 0.61 | 0.86 | 0.95 | 0.23 | 0.85 | 0.72 | 0.90 | 0.59 |
|  | PII | 0.75 | 0.69 | 0.81 | 0.73 | 0.61 | 0.73 | 0.63 | 0.73 | 0.67 |
|  | PIII | 0.71 | 0.60 | 0.79 | 0.74 | 0.61 | 0.71 | 0.69 | 0.72 | 0.67 |
| ANN | PI | 0.63 | 0.54 | 0.73 | 0.97 | 0.26 | 0.90 | 0.57 | 0.94 | 0.62 |
|  | PII | 0.87 | 0.78 | 0.96 | 0.85 | 0.71 | 0.86 | 0.71 | 0.85 | 0.78 |
|  | PIII | 0.89 | 0.75 | 0.99 | 0.93 | 0.77 | 0.91 | 0.84 | 0.92 | 0.85 |
| DT | PI | 0.66 | 0.54 | 0.78 | 0.98 | 0.36 | 0.90 | 0.72 | 0.94 | 0.67 |
|  | PII | 0.83 | 0.73 | 0.92 | 0.88 | 0.66 | 0.84 | 0.74 | 0.86 | 0.77 |
|  | PIII | 0.85 | 0.70 | 0.97 | 0.90 | 0.70 | 0.86 | 0.78 | 0.87 | 0.80 |

Abbreviations: AUC - Area under the receiver operating characteristic curve; AUCL(H) - Lower (Upper) 95% AUC confidence interval calculated for each repetition based on DeLong method<sup>11</sup>; SENS: Sensitivity; SPEC: Specificity; PPV: Positive predictive value, NPV: Negative predictive value; F1: F1 Score; BACC: Balanced accuracy. Highlighted in bold are the values that correspond to the best performing algorithm for each phase.

#### Performance split by indication

To detect whether the performance of the algorithms on the validation set is driven by certain subgroups of projects, we split the validation set based on therapeutic area and report the average AUC and average BACC for each area across ML methods and data sets (Exhibit 22). In line with the results from the complete validation sample, the BACC and AUC values of BART rank highest across therapeutic areas when compared to the other ML approaches. Moreover, we do not find substantial differences between the performance of different indication samples. But the outcome of projects from some indications seem to be predicted more accurate than of others. For example, PII Blood Cancer projects enjoy high classification properties across algorithms while the outcome of PII projects dealing with Cardiovascular diseases seems more challenging to classify correctly. Note we report only therapeutic areas for which occurrences in the validation set are sufficiently frequent (more than 40 projects for PI and PII, more than 30 projects for PIII).

**Exhibit 22:**
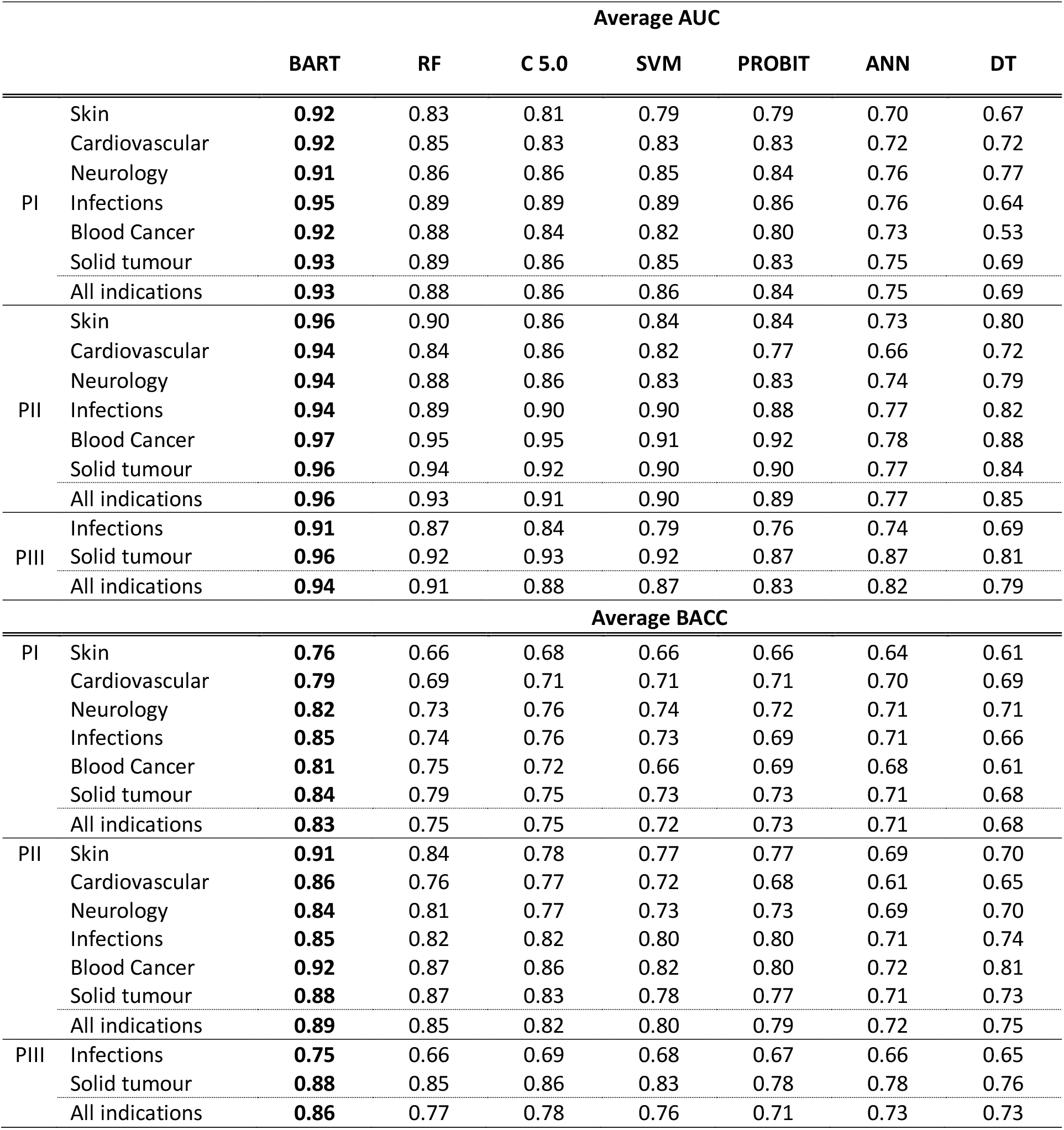
ML performance on validation set split by indication

### 4. Extra graphics and tables

a Projects given expedited FDA review are projects that have received one of the following FDA designations: priority review, breakthrough therapy, accelerated approval, or fast track.

b We pooled projects that reached NDA/BLA with Approved and Marketed ones since it is rare that projects fail during NDA/BLA review. (In our data only 2.6% fail during NDA/BLA review, and 0.1% are not marketed even though approved), which results in too few observations to successfully train ML algorithms.

c Evaluate Ltd. is a commercial company that collects and integrates company-reported and other published pharmaceutical product and financial information to create the EvaluatePharma® database, which includes company pipelines, sales forecasts and proprietary analytics.

d In addition, in Exhibit 18 of the supplementary material A.2, we report the features selected by the backward/forward probabilistic regression used in DISCR together with its coefficients, standard errors and p-values. It provides a notion of the direction of effects and their significance. Note that the selected features overlap with the ones selected by BART, yet DISCR imposes by construction a linear model, not allowing feature interactions.

e The median clinical trial expense was estimated at $15 million for PII and $79 million for PIII.

f Please consult supplementary material A.3 for more details on the robustness checks.

